# ADuLT: An efficient and robust time-to-event GWAS

**DOI:** 10.1101/2022.08.11.22278618

**Authors:** Emil M. Pedersen, Esben Agerbo, Oleguer Plana-Ripoll, Jette Steinbach, Morten Dybdahl Krebs, David M. Hougaard, Thomas Werge, Merete Nordentoft, Anders D. Børglum, Katherine L. Musliner, Andrea Ganna, Andrew J. Schork, Preben B. Mortensen, John J. McGrath, Florian Privé, Bjarni J. Vilhjálmsson

**Author notes:** Contributed equally (Florian Privé, Bjarni J. Vilhjálmsson).

## Abstract

Proportional hazards models have previously been proposed to analyse time-to-event phenotypes in genome-wide association studies(GWAS). While proportional hazards models have many useful applications, their ability to identify genetic associations under different generative models where ascertainment is present in the analysed data is poorly understood. This includes widely used study designs such as case-control and case-cohort designs (e.g. the iPSYCH study design) where cases are commonly ascertained.

Here we examine how recently proposed and computationally efficient Cox regression for GWAS perform under different generative models with and without ascertainment. We also propose the age-dependent liability threshold model (ADuLT), first introduced as the underlying model for the LT-FH++ method, as an alternative approach for time-to-event GWAS. We then benchmark ADuLT with SPACox and standard case-control GWAS using simulated data with varying degrees of ascertainment. We find Cox regression GWAS to underperform when cases are strongly ascertained (cases are oversampled by a factor larger than 5), regardless of the generative model used. In contrast, we found ADuLT to be robust to case-control ascertainment, while being much faster to run. We then used the methods to conduct GWAS for four psychiatric disorders, ADHD, Autism, Depression, and Schizophrenia in the iPSYCH case-cohort sample, which has a strong case-ascertainment. Summarising across all four mental disorders, ADuLT found 20 independent genome-wide significant associations, while case-control GWAS found 17 and SPACox found 8, consistent with our simulation results.

As more genetic data are being linked to electronic health records, robust GWAS methods that can make use of age-of-onset information have the opportunity to increase power in analyses. We find that ADuLT to be a robust time-to-event GWAS method that performs on par with or better than Cox-regression GWAS, both in simulations and real data analyses of four psychiatric disorders. ADuLT has been implemented in an R package called LTFHPlus, and is available on GitHub.

## 1 Introduction

Over the last decade, genome-wide association studies (GWAS) have successfully identified thousands of genetic variants associated with human diseases[18, 54]. Most of these GWASs have modelled the outcome as a binary case-control variable in a logistic (or linear) regression while accounting for covariates such as age, sex, and genetic principal components. However, these models are generally not suited for modelling time-to-event data, as they do not account for certain types of missing or censored data. Time-to-event models are commonly used in epidemiology and many other fields, and have proven useful for both accounting for censoring, changes in disease incidence over time (cohort effects), and age-of-onset[23]. Time-to-event models can also be used to estimate absolute time-dependent risk (i.e. the probability of developing the disease as a function of time) conditional on individual features, and are therefore widely used to estimate disease risk in clinical settings[24].

Although time-to-event models have been proposed for GWAS[19, 49, 37, 48], their adoption has been limited in practice. One reason is that age-of-onset (AOO) information is often not made available. However, time-to-event data is becoming more readily available as more genotyped data are being linked to health records. Another reason is that fitting these models on large data is computationally intensive. However, several computationally efficient survival analysis GWAS methods have been proposed recently for large population-scale data. These include efficient Cox regression implementations[5, 17], and an efficient frailty (random effects) model[11]. The frailty model inherits some of its advantages from the mixed model[53, 22, 30, 32], and can both account for population structure and relatedness, as well as improve statistical power when sample sizes are large. However, to the best of our knowledge, performance of Cox-based regressions in a GWAS setting is limited and they have only been viewed in comparison to other Cox-based regressions or logistic regression[48, 19]. Importantly, these benchmarks have focused on the proportional hazards generative model and without significant case ascertainment, which is common in GWAS. In practice, when collecting data for GWAS it is common to oversample cases to increase the effective sample size and statistical power in the genetic analyses, leading to a case-control or case-cohort study design.

Here we examine to what extent case ascertainment in GWAS data affects Cox regression GWAS and standard case-control GWAS. Inspired by how robust liability threshold models[10, 13] (LTM) have proved to be for ascertained data[55], we propose ADuLT (age-dependent liability threshold) as a computationally efficient time-to-event model for GWAS, and examine how it performs in the presence of case ascertainment. ADuLT is based on the liability threshold model and is the underlying model for the recently proposed LT-FH++ method[1]. ADuLT accounts for age-of-onset information, as well as sex and cohort effects by personalising the thresholds used to infer the case-control status for each individual. These thresholds are personalised by using population-based cumulative incidence proportions (CIPs) for the phenotype of interest as a function of age and additional information, such as sex and birth year (to model sex and cohort effects). We examine how ADuLT compares to SPACox and standard linear regression GWAS in terms of both statistical power and computational efficiency, using both simulations and real iPSYCH data, which is a psychiatric disorder case-cohort data with a strong case ascertainment bias where cases are about 20 times more likely to be sampled[38, 6].

With an increasing integration between biobanks and electronic health records, it is important to utilise additional information in the best way possible, and we believe that knowledge about age-of-onset will be a common and powerful piece of information to include. Finally, ADuLT is implemented in an efficient R package called LTFHPlus (github.com/EmilMiP/LTFHPlus), and is made highly scalable by relying on parallelization and the R package Rcpp, which offers a seamless integration of R and C++[12].

## 2 Methods

### 2.1 Model

The ADuLT model is an extension of the classical LTM[13, 10], and is the model underlying our previously proposed LT-FH++ method[1]. To estimate an individual’s genetic liability, ADuLT utilises birth year, sex, phenotype-specific age-of-onset for cases and current age for controls, as well as population-based cumulative incidences (i.e. the probability of having developed the disease at a given age). In contrast to LT-FH++, the ADuLT model does not incorporate family history as presented here. Instead, we focus on comparing ADuLT to standard time-to-event GWAS methods. ADuLT can account for cohort effects (changes in disease incidence by birth year), as well as differences by sex. This however requires population-based estimates to be available by age, sex and birth year for each phenotype of interest.

The ADuLT model extends the classical LTM by allowing the threshold used to determine case-control status to depend on sex, birth year, and (if available) age-of-onset for an individual. The LTM assumes that each individual has a liability *ℓ* that follows a standard normal distribution in the population. When this liability is larger than a given threshold, *ℓ* ≥ *T*, where *P* (*ℓ* ≥ *T*) = *K* and *K* is the trait’s lifetime prevalence, then the individual is a case (*z* = 1), otherwise it is a control (*z* = 0). This model does not account for time-to-event. Under the ADuLT model, the trait prevalence *K* becomes the available population-based cumulative incidence stratified by sex and birth year, if this information is available. In Figure S12, an example of those CIPs can be seen for depression. Additionally, we assume that the liability can be decomposed into two independent components, a genetic component, *ℓ*_*g*_, and an environmental component, *ℓ*_*e*_, such that *ℓ* = *ℓ*_*g*_ + *ℓ*_*e*_. The genetic liability *ℓ*_*g*_ is normally distributed with mean 0 and variance *h*^2^, where *h*^2^ denotes the trait heritability on the liability scale. The environmental component is normally distributed with mean 0 and variance 1 − *h*^2^ and independent of *ℓ*_*g*_.

ADuLT aims to estimate an expected genetic liability. We do this by expressing the liability as a 2-dimensional normal distribution given by:

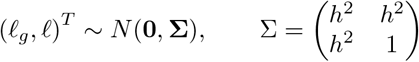

The mean of the genetic component is given by

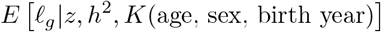

where the information we condition on, namely the case-control status and CIPs, result in an interval of (full) liabilities to integrate over. The CIPs set the threshold and the case status determines if the integration is above (a case) or below (control) the threshold.

### 2.2 Simulation Details

The default simulation setup uses two generative models, namely the Cox proportional hazards model and the LTM. We simulate under both generative models in order not to favour one method over the other.

Initially, genotypes are simulated for *N* = 1, 000, 000 individuals and *M* = 20, 000 independent SNPs. The genotypes are sampled from a binomial distribution *Binom*(2, *AF*) with the probability parameter set to the allele frequency (AF) of a given SNP. The AFs are sampled from a uniform distribution on the interval (0.01, 0.49). SNPs are standardised using the true AF, and for the scaled SNPs, the effect sizes of causal SNPs were drawn from the normal distribution *N* (0, *h*^2^*/C*), where *C* denotes the number of causal SNPs and *h*^2^ denotes the liability-scale heritability. In the simulations, we used *h*^2^ = 0.5 and either *C* = 250 or *C* = 1000 causal SNPs. With the simulated genotypes and causal effect sizes, we then assigned synthetic phenotypes using the two generative models.

For the proportional hazards model, we opted for a simulation setup as similar as possible to the one used in SPACox[5]. First, we simulated the censoring times, *c*_*i*_, for each individual *i* from an exponential distribution with a scale parameter of 0.15. Next, we simulated onset times[4], 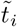, using a Weibull distribution[2] as follows

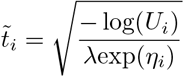

where *λ* is the event rate, *U*_*i*_ ∼ Unif(0, 1), 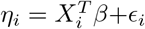, with *ϵ*_*i*_ ∼ *N* (0, 1 − *h*^2^), and 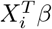 are the scaled genotypes multiplied by effect sizes, corresponding to the genetic liability *ℓ*_*g*_ in the LTM. The case-control status *z*_*i*_ is then 1 if 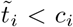, and 0 otherwise. The event time 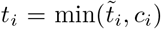 is the observed time. The event rate *λ* was chosen such that the lifetime prevalence is fixed at e.g. 1% or 5%. The simulation of onset times depends on all causal SNPs, which deviates from the simulations of onset times in the SPACox paper, where the onset times depended on a single causal SNP only. This change was made in order for the full genetic load of an individual to influence the onset times, instead of just a single SNP. Next, we calculated the CIP of the simulated event times, i.e. the fraction of cases observed before a given point in time. then the proportions were converted to the ages-of-onset (in years) using the logistic function given by Equation (1) with median age-of-onset *x*_0_ = 50 and growth rate *k* = 0.2. Both age and age-of-onset were used to calculate the cumulative incidence proportions, which in turn defines the thresholds under the ADuLT model. For instance, with a lifetime prevalence of 1%, 90% of all individuals had an age or age-of-onset between 17 and 57 years.

Under the LTM, we set the trait status *z*_*i*_ equal to 1 if the liability exceeds the threshold, i.e. if *ℓ*_*i*_ *> T*, and 0 otherwise, where 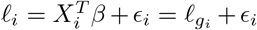. The threshold *T* is determined by the lifetime prevalence *K*. For instance, a lifetime prevalence of 5% and 10% results in thresholds *T* = 1.64 and *T* = 1.28, respectively. The relationship between the age-of-onset and the liabilities above the threshold *T*, is given by the logistic function

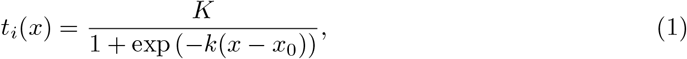

where *K* denotes the maximal attainable value, *k* denotes the growth rate, and *x*_0_ denotes the median age-of-onset. Using the age of controls, we know how long they have lived without being diagnosed. This information allows us to exclude liabilities, i.e. the period of risk lived through so far. For both cases and controls, the personalised thresholds are calculated as *T*_*i*_ = Φ(1 − *CIP*_*i*_), where *T*_*i*_ is the personalised threshold and *CIP*_*i*_ is the CIP for individual *i*. The liabilities below the personalised threshold are considered for controls and the liabilities above the threshold are considered for cases. If the population-representative CIPs are stratified by birth year and sex, the full liaiblity for cases can be fixed at *T*_*i*_. Ages for controls are sampled from a uniform distribution between 10 and 90. This resulted in 90% of individuals having an age between 14 and 86.

### 2.3 The ADuLT survival model

As we showed previously[1], the age-dependent liability threshold model can be considered a survival model. More specifically, consider the survival function *S*_*i*_(*age*) = *P* (age_*i*_ *> age*), where age_*i*_ represents age-on-set for cases or censoring time for the *i*^*th*^ individual. The probability that an individual has not become a case for a given *age* is equal to the probability that the individual’s liability is larger than the (individualised) liability threshold *T*_*i*_(*age*), which is a shorthand notation for the age-dependent threshold given by

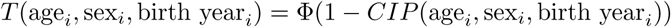

. Here sex_*i*_ and birth year_*i*_ are the *i*^*th*^ individual’s sex and birth year, respectively. If we assume that the individual liability consists of a genetic and an environmental component, 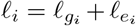, where 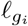 and 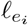 are Gaussian distributed with mean 0 and variance *h*^2^ and 1 − *h*^2^, respectively, then we can write the survival function as follows

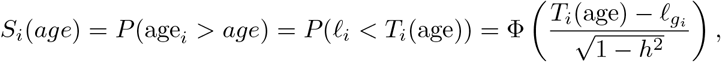

where Φ is the standard Gaussian cumulative distribution function and we assume that the genetic liability contribution is known. In the last equality, we standardise the environmental contribution with the known genetic contribution and the variance. From this we can derive the event density, and the hazard function for the *i*^*th*^ individual as

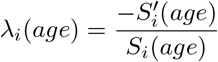

We note that this survival model is unusual in a couple of ways. First, each individual has a slightly different parameterisation of the model, which comes through the individualised liability threshold *T*_*i*_(*age*). Second, the genetic effects affect the hazard rate by shifting the individual liability. Third, *T*_*i*_(*age*) does not have to approach negative infinity as *age* approaches positive infinity, but may instead simply become fixed for all values *T*_*i*_(*age*) above some threshold, e.g. if every individual in a cohort has died and no new event are possible. This is not necessarily a problem for the interpretation as *T*_*i*_(*age*) may still be piece-wise differentiable, and the hazard rate for all values t above this threshold then becomes 0.

### 2.4 GWAS in iPSYCH

With the second wave of genotyped individuals, the iPSYCH case-cohort reached ∼143,000 individuals, up from ∼80,000.[6] Both waves have been imputed with the RICOPILI imputation pipeline[26], and were then combined into a single dataset. We restricted the analysis to SNPs that passed RICOPILI quality controls for both waves, resulting in a total of 8,785,478 SNPs for the GWAS. The analysis was restricted to a group of individuals with European ancestry, which were identified by calculating a robust Mahalanobis distance based on the first 20 PCs and restricting to a log-distance below 4.5[43]. We filtered for relatedness by removing individuals (the second one in each pair) with a KING-relatedness above 0.088. Since the iPSYCH case-cohort has a population representative subcohort and oversampled cases for six major psychiatric disorders (here we focus on ADHD, autism, depression and schizophrenia), we restricted each analysis to the individuals in the subcohort (which is a random sample of the entire population) and the cases for the phenotype being analysed, i.e. oversampled cases from the other psychiatric disorders were not used. The final number of individuals used for the GWAS of each phenotype is presented in Table S2. The linear regression GWAS was performed using the bigsnpr package[41] for R and SPACox GWAS was performed using the original implementation in the SPACox package for R. We used 20 PCs, sex, and imputation wave as covariates for all analyses. We included age as a covariate when analysing case-control status. Age was not included as a covariate when using the ADuLT phenotype or SPACox. We chose not to use a mixed model approach for GWAS with case-control status or ADuLT phenotypes, as SPACox did not have a similar option for random effects.

### 2.5 Cumulative Incidence Proportions

The CIPs can be interpreted as the proportion of individuals diagnosed with a certain disorder before a given age. As a result, the CIPs are population and disorder specific and can be stratified by sex and birth year. The CIPs used here were stratified by sex and birth year to account for differences in incidences between sexes and for different birth years (cohorts). The CIPs were estimated from Danish population-based registers. The Danish Civil Registration System[39] was used to identify individuals and contains all 9,251,071 individuals that lived in Denmark at some point between April 2, 1968 and December 31, 2016. The Danish Civil Registration System has continually recorded information since its launch in 1968, and includes information about sex, date of birth, date of death, and date of emigration, or immigration. Each individual has a unique identifier that can be used to link information of several registers. Information on psychiatric disorders was obtained from the Danish Psychiatric Central Research Register[33]. It contains all admissions to psychiatric inpatient facilities since 1969 and visits to outpatient psychiatric departments and emergency departments since 1995. From 1969 to 1993, the International Classification of Diseases, eighth revision (ICD-8) was used as the diagnostic system. From 1994 onwards, the tenth revision (ICD-10) was used. The four disorders of interest were identified by the following ICD-8 and ICD-10 codes: ADHD (308.01 and F90.0), autism (299.00, 299.01, 299.02, 299.03 and F84.0, F84.1, F84.5, F84.8, F84.9), depression (296.09, 296.29, 298.09, 300.49 and F32, F33), and schizophrenia (295.x9 excluding 295.79 and F20). The age-of-onset was defined as the age of an individual at first contact with the psychiatric care system, either inpatient, outpatient, or emergency visits. In the analyses, each individual was followed from birth, immigration, or January 1, 1969 (whichever happened last) until death, emigration, or December 31, 2016 (whichever happened first). The cumulative incidence function was estimated separately for each sex and birth year, and the Aalen-Johansen approach was used with death and emigration as competing events[15].

## 3 Results

### 3.1 Overview of method

The age-dependent liability threshold model presented here was first introduced in our previous paper extending the LT-FH method to account for family history as well as age-of-onset, sex, and cohort effects among all individuals, including the family members[21, 1]. In this paper, we focus on the ADuLT model as an alternative to commonly used time-to-event or linear regression GWAS methods, without considering any family history.

The ADuLT model modifies the LTM by assuming that the threshold used to determine an individual’s case-control status corresponds to the CIP at the age of diagnosis. In Figure 1, we present the CIPs for ADHD for individuals born in Denmark in the year 2000. The CIPs increase as the population gets older, which in turn leads to a decreased threshold. If additional information, such as sex and birth year, is available, the population CIPs should be stratified according to this additional information (as seen in Figure 1), as this improves estimation of the genetic liability[1]. In the first step, a personalised threshold is assigned to each individual based on their current age or the age-of-onset, as well as sex and birth year. In the second step, the ADuLT model uses the liability-scale heritability to estimate a genetic liability for each individual. The third step uses the ADuLT phenotype as a continuous outcome in a GWAS. There are no restrictions on the choice of GWAS method as long as it accepts continuous outcomes, allowing researchers to benefit from current and future advances in GWAS methods. Note that Figure 1 illustrates the use of CIP for cases. If an individual is a control, the area of possible liabilities will instead be from negative infinity to the threshold identified from the CIPs.

**Figure 1:**
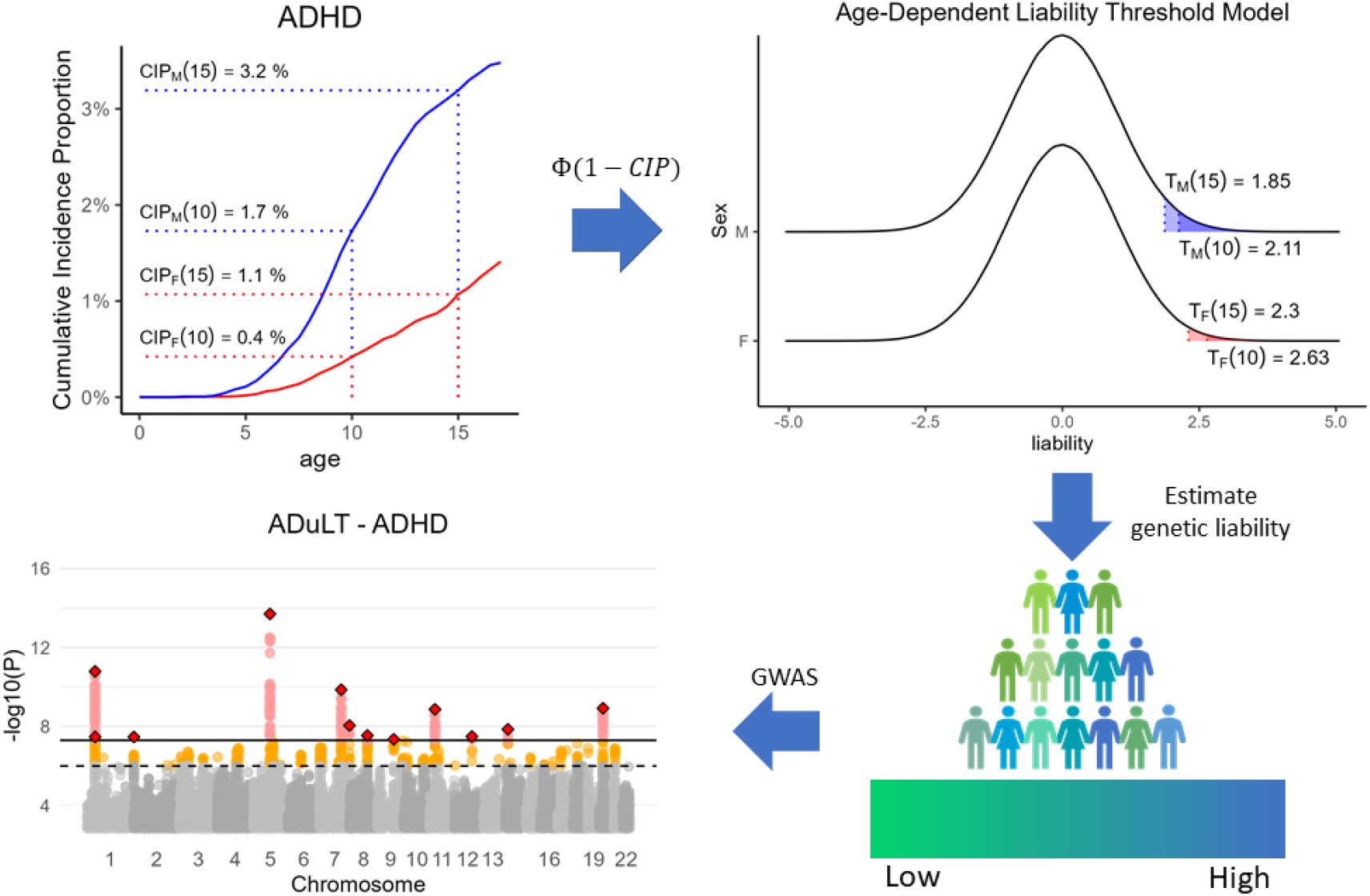
Overview of ADuLT and illustration of the information it can account for. Overview of the information used, and the different steps needed to perform a GWAS based on the ADuLT phenotype. The cumulative incidence proportions (CIPs) stratified by sex and birth year (here ADHD for individuals born in Denmark in 2000) are converted to a threshold for the age-dependent liability threshold model. Females are represented by the red line, while males are represented by the blue line. The CIPs has been marked at the age of 10 and 15 for both sexes (dotted lines). Finally, a genetic liability is estimated for each individual, and this ADuLT phenotype can be used as the outcome in a GWAS.

### 3.2 Simulation Results

We used two generative models for the simulations, namely the LTM and the proportional hazards model (see Methods). The performance of a simple case-control GWAS, SPACox, and the ADuLT phenotype used as the outcome in a linear regression-based GWAS was assessed under both generative models. Sex and age or age-of-onset were simulated for 1 million individuals, each with 20,000 independent SNPs. To examine the effect of ascertainment of cases, which is common in GWAS data, similar analyses were performed where the total number of individuals was randomly downsampling from 1 million to 20,000 individuals, leaving 10,000 controls and 10,000 cases in each downsampled dataset.

Figure 2 displays the power for each method under both generative models with 250 causal SNPs. A similar plot showing the power of the same generative models but with 1000 causal SNPs can be found in Figure S1. Without downsampling, the power of all three methods is similar under both generative models (Figure 2A). In Figure 2B, which is based on a downsampled data set simulating case ascertainment, the power of all three methods decreased due to a reduced sample size, but the power of SPACox was disproportionately affected by the downsampling. For simulated traits that have been downsampled and have a lifetime prevalence of 5% or below, SPACox performs worse than linear regression for both the case-control status and the ADuLT phenotype by more than a factor of 10 in the worst case, and approximately 25% worse in the best case. Under the proportional hazards model and a lifetime prevalence of 20%, and with downsampling, SPACox has an average power on par with ADuLT.

**Figure 2:**
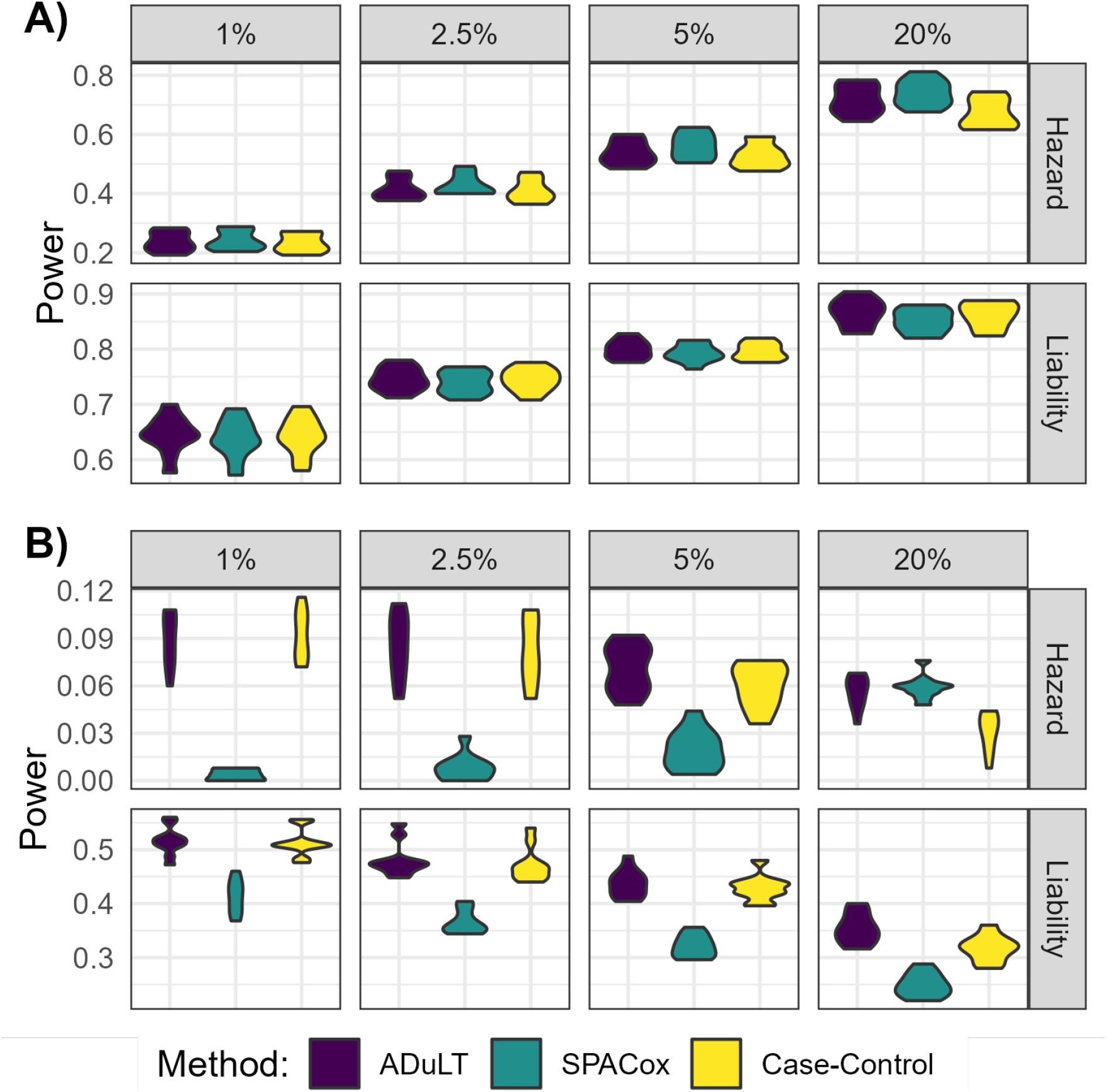
Power simulation results with 250 causal SNPs under both generative models and varying prevalences. The power is shown for different population prevalence, varying from 1% to 20%. **A)** The power, i.e. the fraction of causal SNPs detected for each method, **without downsampling. B)** The power **with downsampling**, i.e. the number of individuals is subsampled to 10k cases and 10k controls.

In Table S1, which is based on data simulated under the LTM and without downsampling, the relative power of all methods are within 3% of one another. ADuLT obtained the highest power in all cases, while the lowest power was observed in connection to SPACox. With downsampling and 1000 causal SNPs, the increase in power was 117% with ADuLT over SPACox across all prevalences considered, and it was 96% for case-control status over SPACox. With downsampling and 250 causal SNPs, we observed an average increase of 34% in power over SPACox with ADuLT, and a 29% increase in power with case-control status, showing that SPACox has a comparatively low power for low effect sizes.

In Figures S2 and S3, the average *χ*^2^-statistics for the null SNPs is reported. Plots were achieved for 250 and 1000 causal SNPs, respectively, and each plot contains results for four different lifetime prevalences, with and without downsampling, and for both generative models. All models were well calibrated, since no inflation of the null statistics is observed. Figures S4 and S5 show the relative power with SPACox as the baseline method. As before each plot holds the relative power for four lifetime prevalences, with and without downsampling and both generative models. In addition, different plots were achieved for 250 and 1000 causal SNPs. For 250 causal SNPs and no downsampling, performance of all methods were similar. However, with downsampling, SPACox only identified a few causal SNPs, which resulted in large relative power gains for ADuLT and linear regression (see Figure 2B). In Table S1, simulation results for all parameter setups are available, including the power, relative power compared to SPACox, and mean chi-square statistic of null SNPs.

#### 3.2.1 Computation Times

The computational time for estimating the ADuLT phenotype depends solely on the number of individuals. The running time for the GWAS step depends heavily on the implementation of the GWAS method used. In Figure 3, the combined running times of estimating the ADuLT phenotype and performing a GWAS using the bigsnpr package[41] are reported. We used 4 CPU cores for both steps, which is a conservative number of cores. The SPACox implementation does not support parallelization, which is why SPACox was run sequentially. We find that ADuLT together with a linear regression is faster than SPACox, even with only modest parallelization. Logistic regression of a binary phenotype is slower than linear regression of the same phenotype[41], which means ADuLT together with a linear regression may be faster and have higher power to detect causal SNPs.

**Figure 3:**
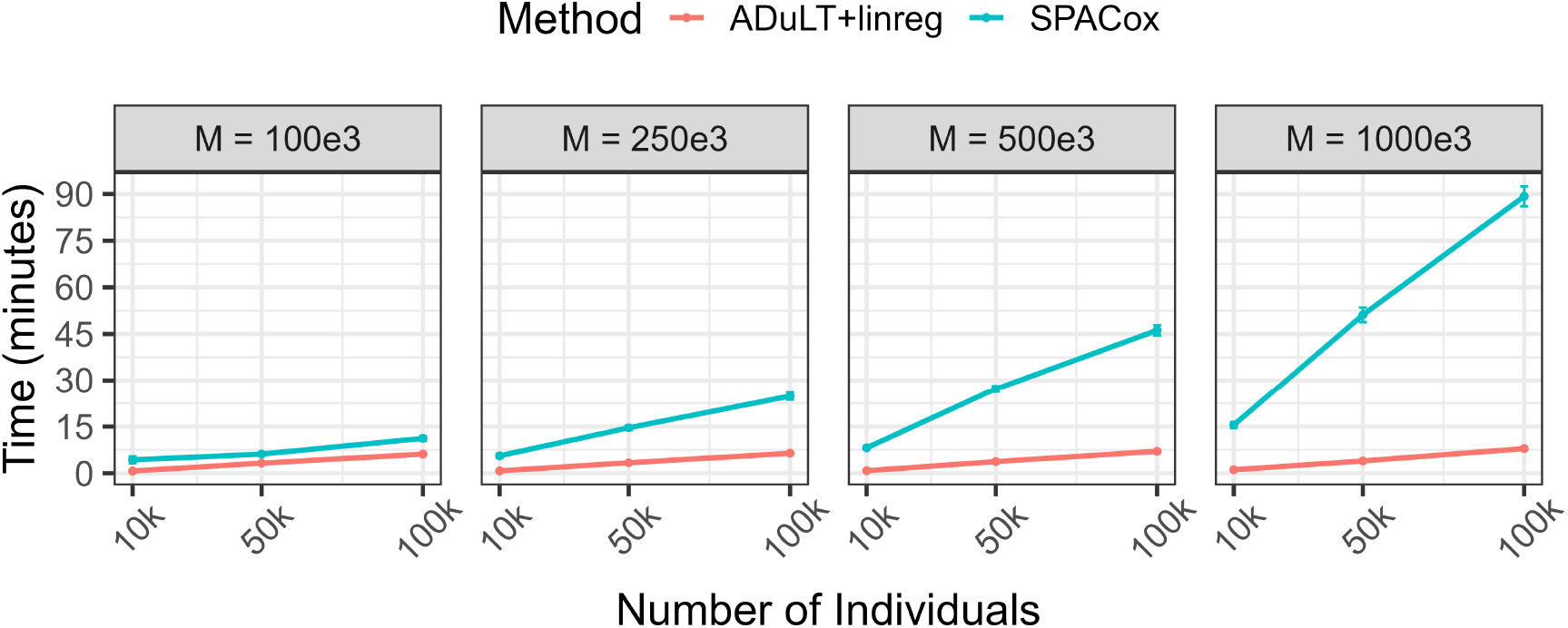
Running times of ADuLT combined with a linear regression GWAS compared to SPACox. Each point represents the mean value of 10 replications, while the error bars are represented by the estimate ±1.96 standard errors. Run times were assessed for a varying number of individuals and SNPs (M). SPACox uses a single CPU core, as no parallelization is available. We used 4 CPU cores for estimating the ADuLT phenotype and performing the linear regression GWAS for this phenotype. The means and corresponding standard errors of the run times can be found in Table S1.

### 3.3 GWAS of psychiatric disorders in iPSYCH

The iPSYCH data has been linked to the Danish registers, which means that detailed information on age-of-onset, age, sex, and birth year can be assessed for all genotyped individuals that are part of the iPSYCH cohort[6] This supplementary information was used to analyse four psychiatric traits, namely ADHD, autism, depression, and schizophrenia. For each of these phenotypes, population-based CIPs were obtained by birth year and sex (see Figures S6, S9, S12 and S15 for plots of the CIPs used, and see Cumulative Incidence Porportions for details). The prevalences were used to tailor the thresholds to each individual under the ADuLT model (see Methods).

We performed GWASs for each of the four phenotypes and for each of the methods considered, i.e. using either the case-control status or the estimated genetic liability by ADuLT as the outcome in a linear regression-based GWAS or SPACox (see Methods for details). Figure 4 displays the Manhattan plots for ADHD for all methods, where the case-control GWAS included age as a covariate, while the ADuLT GWAS and SPACox did not. To report nearly independent findings, LD clumping was performed on the summary statistics with a *r*^2^ threshold of 0.1 and a window size of 500kb, prioritising the SNPs with the lowest p-values. This was done for each combination of phenotype and method. The lowest p-value LD-clumped SNPs that are unique to ADuLT and ADHD can be found in Table S3 and the LD-clumped snps that are unique to case-control status and ADHD can be found in Table S4. For ADHD, we found 12 independent genome-wide significant associations when using the ADuLT phenotype as the outcome, while case-control status and SPACox found 11 and 5 associations, respectively. The ADuLT GWAS had two independent associations that were not identified by case-control associations, and case-control GWAS found one association that was not found by the ADuLT GWAS. One of the associations unique to ADuLT is rs4660756. The gene closest to this SNP is ST3GAL3, which has previously been associated with educational attainment[36] and ADHD[52]. SPACox also identified ST3GAL3, but through rs11810109 instead. The association unique to case-control GWAS is rs8085882 on chromosome 18. The closest gene is ZNF521, which has previously been associated with education attainment[29], ADHD[46], and smoking initiation[27]. The association with the lowest p-value that is shared among all methods is rs4916723 on chromosome 5 with LINC00461 as the closest gene. This gene has also been reported as being associated with educational attainment[27] and ADHD[9].

**Figure 4:**
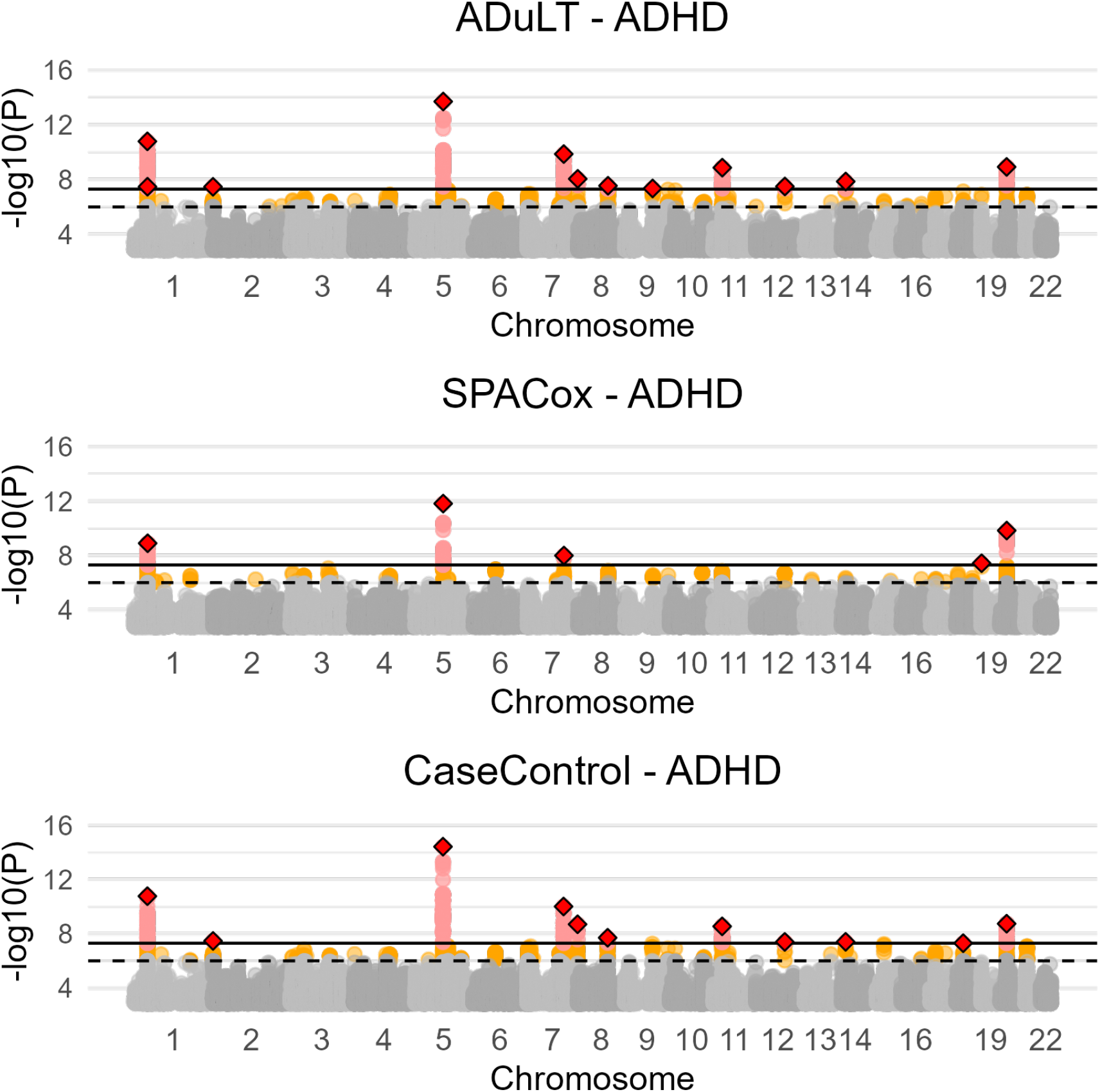
Manhattan plots from GWAS with the ADuLT phenotype, SPACox, and case-control status for ADHD. Manhattan plots for ADHD for all three methods. Case-control GWAS uses the age of individuals as a covariate, whereas the ADuLT GWAS and SPACox do not. The orange dots indicate suggestive SNPs with a p-value threshold of 5 × 10^−6^. The red dots correspond to genome-wide significant SNPs with a p-value threshold of 5 × 10^−8^. The diamonds correspond to the lowest p-value LD clumped SNP in a 500k base pair window with an *r*^2^ = 0.1 threshold.

Across the four psychiatric disorders, ADuLT found 20 independent genome-wide significant associations, while case-control status found 17 and SPACox found 8. The Manhattan plots for each of the methods, each of the remaining disorders (autism, depression, and schizophrenia), and with and without age as a covariate can be found in Figures S7, S8, S10, S11, S13, S14, S16 and S17. Notably, SPACox consistently identified fewer associations than the ADuLT and case-control status GWASs, and was the only method that did not identify any significant association for major depression and schizophrenia.

## 4 Discussion

With biobanks such as the UK biobank[7], iPSYCH[6], FinnGen[25], or Biobank Japan[34] linking electronic health records to genetic data, there is an increased incentive to develop methods that can fully utilise this supplementary information. This includes details about age-of-onset, which can be used in time-to-event analyses to improve power. In epidemiology, time-to-event analyses are usually performed with a Cox-based regression, whereas time-to-event GWAS are still relatively uncommon. This has in part been due to computational challenges of applying Cox regression to GWAS, but recent developments of efficient Cox-based regression methods such as SPACox or GATE have largely resolved this limitation[5, 11]. However, the performance of Cox-based regressions for GWAS has only been viewed in comparison to other Cox-based or logistic regression[48, 19], and not when the case-control cohort is sampled with ascertainment (e.g. where cases are oversampled). Evaluating their performance in ascertained case-control cohorts is important as such datasets are very common in genetics, e.g. the iPSYCH and FinnGen data.

In this paper, we have examined the proportional hazards model implemented in SPACox and found that in situations where cases are ascertained or oversampled (which is often the case in GWAS datasets), the proportional hazards based model was less powerful than a simple linear regression. We proposed the age-dependent liability threshold (ADuLT) model as an efficient and robust alternative to Cox-based time-to-event GWAS. The ADuLT model is the model underlying the recently published LT-FH++ method[1], as presented here it does not incorporate information on family members. However, the main focus of this paper was to compare the ADuLT model to a computationally efficient time-to-event GWAS method, SPACox, without accounting for information such as family history. ADuLT incorporates time-to-event information into the LTM by using liability thresholds that vary with age and sex. These personalised thresholds are derived from population-based estimates of the cumulative incidence proportions. Using this information, ADuLT first estimates individual posterior genetic liabilities, which are then used as a quantitative phenotype in GWAS. This final step can be performed with any GWAS software, which allows for ADuLT to benefit from using advanced GWAS methods, such as linear mixed models[30, 22, 32]. The computational cost of estimating the individual posterior liabilities is neglectable when compared to the computational cost of performing even a simple GWAS with linear regression.

Using simulations, we compared different GWAS methods, Cox regression as implemented in SPACox and a linear regression with the ADuLT phenotype and the case-control status. As expected we found a Cox-based time-to-event GWAS to provide most power under the proportional hazards generative model, however it was closely followed by the ADuLT GWAS and case-control GWAS, especially when disease prevalence is low. Conversely, when simulating under the LTM, the ADuLT GWAS had the greatest power, followed by Cox regression and case-control GWAS. However, when considering ascertainment of cases, we found SPACox to have the lowest power of all considered methods under both generative models and for all prevalences except one (the least ascertained sample). We note that these results are in line with previously reported comparison between Cox regression and linear regression in case-cohort studies[48]. When we applied all three methods to the iPSYCH data, which has a high degree of case ascertainment, the results were in agreement with the simulation results in that SPACox identified fewer genome-wide significant variants than the case-control or the ADuLT GWASs. Therefore, for identifying significant genome-wide associated variants, a Cox-regression GWAS can have less statistical power than linear regression with case-control status or the ADuLT phenotype. As a result, we recommend using more robust GWAS methods, such as on case-control status or the ADuLT phenotype when performing GWAS in ascertained samples, which includes most case-cohort and case-control datasets.

Although Cox regression GWAS may not be robust to ascertained samples, we note that it can still improve power in population cohorts, and may still yield unbiased estimates. Furthermore, several adjustments have been proposed to Cox regression when applied to ascertained data, such as inverse probability weighting[45] (IPW). IPW results in unbiased estimates, but estimating their variance (and association p-values) can be difficult[3]. Furthermore, to the best of our knowledge, IPW is currently not implemented in computationally efficient Cox regression GWAS methods (e.g. SPACox).

In contrast, we found ADuLT to be a computationally efficient and robust approach for time-to-event GWAS. Moreover, using the LTM, it is possible to account for family history information[1, 21], and it can be used in connection to risk prediction[20, 8, 47]. GWAS individual-level data can also be used to build polygenic scores based on efficient penalized regression models[42]; a future direction of research for us is to investigate whether a penalized linear regression using the ADuLT-inferred outcome would be preferable to using a Cox-based penalized regression as implemented in e.g. snpnet-Cox[28]. As other possible future directions, the ADuLT model may also provide an alternative framework for examining interactions between age and genetic variants[35], and provide insight into the genetics underlying disease trajectory. Like LT-FH[21] and LT-FH++[1], ADuLT also has the advantage that it produces quantitative posterior liabilities which can be treated as quantitative phenotypes and analysed with advanced GWAS method, such as BOLT-LMM[30], fastGWA[22], or REGENIE[32]. However, ADuLT does have some limitations. First, it requires population-representative CIPs to be available for the disorder of interest, and preferably stratified by sex and birth year. Recent efforts to make such data publicly available for all diseases is therefore of great interest[40]. Second, the assumption that early onset cases have higher disease liability may not always be true. Although age-of-onset tends to be negatively genetically correlated with case-control status, the correlation is not always very strong[14]. Third, the model does not account for possible interactions between genotype (or environment) with age, but exploring methods that model this relationship is a future direction. Fourth, similar to LT-FH[21] and LT-FH++[1], ADuLT assumes the narrow sense (additive) heritability is known a priori for the outcome of interest. These can either be obtained from literature or estimated in the data, e.g. using family-based heritability estimates[56]. However, we have also previously shown that the model we use is robust to misspecification of prevalence information and heritability[1]. Finally, in this study we did not consider downsampling of cases or ascertainment of healthy controls, which might be relevant for many genetic datasets such as the UK biobank[7] or the Danish blood donor study data[16].

As age information becomes more readily available, we expect time-to-event methods for GWAS that make use of such information to become more common. However, the benefit of these methods may depend on how the data was collected, as well as their ability to account for other confounders. We believe ADuLT provides both a robust, computationally efficient, and a flexible approach for time-to-event analyses in population-scale datasets.

## Data Availability

iPSYCH is approved by the Danish Scientific Ethics Committee, the Danish Health Data Authority, the Danish Data Protection Agency, Statistics Denmark, and the Danish Neonatal Screening Biobank Steering Committee. Code used to generate simulation results, analyse iPSYCH, and generate plots and tables can be found at https://github.com/EmilMiP/ADuLTCode. LT-FH++ can be found at https://github.com/EmilMiP/LTFHPlus.

## 5 Data and code availability

iPSYCH is approved by the Danish Scientific Ethics Committee, the Danish Health Data Authority, the Danish Data Protection Agency, Statistics Denmark, and the Danish Neonatal Screening Biobank Steering Committee[38]. Code used to generate simulation results, analyse iPSYCH, and generate plots and tables can be found at https://github.com/EmilMiP/ADuLTCode. LT-FH++ can be found at https://github.com/EmilMiP/LTFHPlus.

## 6 Acknowledgements

We would like to thank Jakob Grove, Doug Speed, Matthew Robinson, and Sven Erik Ojavee for valuable discussions. E.M.P, B.J.V. and F.P. were supported by the Lundbeck Foundation Initiative for Integrative Psychiatric Research, iPSYCH (R102-A9118, R155-2014-1724 and R248-2017-2003), and a Lundbeck Foundation Fellowship (R335-2019-2339). J.M., B.J.V. and F.P. were also supported the Danish National Research Foundation (Niels Bohr Professorship to Prof. John McGrath). A.J.S. is supported by a Lundbeckfonden Fellowship (R335-2019-2318), and O.P.R. is supported by a Lundbeck Foundation Fellowship (R345-2020-1588). K.M. is supported by grants from The Lundbeck Foundation (R303-2018-3551) and the Brain & Behavior Research Foundation (Young Investigator Award 2021). A.G. has received funding from the European Research Council (ERC) under the European Union’s Horizon 2020 research and innovation programme (grant agreement No 945733), starting grant AI-Prevent. High-performance computer capacity for handling and statistical analysis of iPSYCH data on the GenomeDK HPC facility was provided by the Center for Genomics and Personalised Medicine and the Centre for Integrative Sequencing, iSEQ, Aarhus University, Denmark (grant to A.D.B.).

## References

[1] “Accounting for age of onset and family history improves power in genome-wide association studies”. In: Am. J. Hum. Genet. (Feb. 2022).

[2] Peter C Austin. “Generating survival times to simulate Cox proportional hazards models with time-varying covariates”. en. In: Stat. Med. 31.29 (Dec. 2012), pp. 3946–3958.

[3] Peter C Austin. “Variance estimation when using inverse probability of treatment weighting (IPTW) with survival analysis”. en. In: Stat. Med. 35.30 (Dec. 2016), pp. 5642–5655.

[4] Ralf Bender, Thomas Augustin, and Maria Blettner. Generating survival times to simulate Cox proportional hazards models. 2005.

[5] Wenjian Bi et al. “A Fast and Accurate Method for Genome-Wide Time-to-Event Data Analysis and Its Application to UK Biobank”. en. In: Am. J. Hum. Genet. 107.2 (Aug. 2020), pp. 222–233.

[6] Jonas Bybjerg-Grauholm et al. “The iPSYCH2015 Case-Cohort sample: updated directions for unravelling genetic and environmental architectures of severe mental disorders”. en. In: medRxiv (Dec. 2020), p. 2020.11.30.20237768.

[7] Clare Bycroft et al. “The UK Biobank resource with deep phenotyping and genomic data”. en. In: Nature 562.7726 (Oct. 2018), pp. 203–209.

[8] Shai Carmi. “Cascade screening following a polygenic risk score test: what is the risk of a relative conditional on a high score of a proband?” In: bioRxiv (2021).

[9] Ditte Demontis et al. “Discovery of the first genome-wide significant risk loci for attention deficit/hyperactivity disorder”. en. In: Nat. Genet. 51.1 (Nov. 2018), pp. 63–75.

[10] E R Dempster and I M Lerner. “Heritability of Threshold Characters”. en. In: Genetics 35.2 (Mar. 1950), pp. 212–236.

[11] Rounak Dey et al. “An efficient and accurate frailty model approach for genome-wide survival association analysis controlling for population structure and relatedness in large-scale biobanks”. en. Nov. 2020.

[12] Dirk Eddelbuettel and Romain Francois. “Rcpp: Seamless R and C++ Integration”. en. In: J. Stat. Softw. 40 (Apr. 2011), pp. 1–18.

[13] D S Falconer. “The inheritance of liability to certain diseases, estimated from the incidence among relatives”. In: Ann. Hum. Genet. 29.1 (Aug. 1965), pp. 51–76.

[14] Yen-Chen A Feng et al. “Findings and insights from the genetic investigation of age of first reported occurrence for complex disorders in the UK Biobank and FinnGen”. Nov. 2020.

[15] Stefan N Hansen et al. “Estimating a population cumulative incidence under calendar time trends”. en. In: BMC Med. Res. Methodol. 17.1 (Jan. 2017), pp. 1–10.

[16] Thomas Folkmann Hansen et al. “DBDS Genomic Cohort, a prospective and comprehensive resource for integrative and temporal analysis of genetic, environmental and lifestyle factors affecting health of blood donors”. en. In: BMJ Open 9.6 (June 2019), e028401.

[17] Liang He and Alexander M Kulminski. “Fast Algorithms for Conducting Large-Scale GWAS of Age-at-Onset Traits Using Cox Mixed-Effects Models”. en. In: Genetics 215.1 (May 2020), pp. 41–58.

[18] David M Howard et al. “Genome-wide meta-analysis of depression identifies 102 independent variants and highlights the importance of the prefrontal brain regions”. en. In: Nat. Neurosci. 22.3 (Feb. 2019), pp. 343–352.

[19] Jacob J Hughey et al. “Cox regression increases power to detect genotype-phenotype associations in genomic studies using the electronic health record”. en. In: BMC Genomics 20.1 (Nov. 2019), p. 805.

[20] Margaux L A Hujoel et al. “Incorporating family history of disease improves polygenic risk scores in diverse populations”. en. Apr. 2021.

[21] Margaux L A Hujoel et al. “Liability threshold modeling of case-control status and family history of disease increases association power”. en. In: Nat. Genet. 52.5 (May 2020), pp. 541–547.

[22] Longda Jiang et al. “A resource-efficient tool for mixed model association analysis of large-scale data”. en. In: Nat. Genet. 51.12 (Nov. 2019), pp. 1749–1755.

[23] Per Kragh and Niels Andersen. Survival Analysis, Overview. John Wiley & Sons, Ltd, 2014.

[24] Per Kragh Andersen et al. “Analysis of time-to-event for observational studies: Guidance to the use of intensity models”. In: JOUR (2021).

[25] Mitja I Kurki et al. “FinnGen: Unique genetic insights from combining isolated population and national health register data”. en. In: medRxiv (Mar. 2022), p. 2022.03.03.22271360.

[26] Max Lam et al. “RICOPILI: Rapid Imputation for COnsortias PIpeLIne”. en. In: Bioinformatics 36.3 (Feb. 2020), pp. 930–933.

[27] J J Lee et al. “Gene discovery and polygenic prediction from a genome-wide association study of educational attainment in 1.1 million individuals”. In: Nat. Genet. 50.8 (July 2018).

[28] Ruilin Li et al. “Fast Lasso method for large-scale and ultrahigh-dimensional Cox model with applications to UK Biobank”. In: Biostatistics 23.2 (2022), pp. 522–540.

[29] M Liu et al. “Association studies of up to 1.2 million individuals yield new insights into the genetic etiology of tobacco and alcohol use”. In: Nat. Genet. 51.2 (Feb. 2019).

[30] Po-Ru Loh et al. “Efficient Bayesian mixed-model analysis increases association power in large cohorts”. en. In: Nat. Genet. 47.3 (Feb. 2015), pp. 284–290.

[31] A Mahajan et al. “Fine-mapping type 2 diabetes loci to single-variant resolution using high-density imputation and islet-specific epigenome maps”. In: Nat. Genet. 50.11 (Nov. 2018).

[32] Joelle Mbatchou et al. “Computationally efficient whole-genome regression for quantitative and binary traits”. en. In: Nat. Genet. 53.7 (May 2021), pp. 1097–1103.

[33] Ole Mors, Gurli P Perto, and Preben Bo Mortensen. “The Danish Psychiatric Central Research Register”. en. In: Scand. J. Public Health 39.7 Suppl (July 2011), pp. 54–57.

[34] A Nagai et al. “Overview of the BioBank Japan Project: Study design and profile”. In: Journal of epidemiology 27.3S (Mar. 2017).

[35] Sven E Ojavee et al. “Novel discoveries and enhanced genomic prediction from modelling genetic risk of cancer age-at-onset”. Mar. 2022.

[36] A Okbay et al. “Genome-wide association study identifies 74 loci associated with educational attainment”. In: Nature 533.7604 (May 2016).

[37] Kouros Owzar et al. “Power and sample size calculations for SNP association studies with censored time-to-event outcomes”. en. In: Genet. Epidemiol. 36.6 (Sept. 2012), pp. 538–548.

[38] C B Pedersen et al. “The iPSYCH2012 case–cohort sample: new directions for unravelling genetic and environmental architectures of severe mental disorders”. In: Mol. Psychiatry 23 (Sept. 2017), p. 6.

[39] Carsten Bøcker Pedersen. The Danish Civil Registration System. 2011.

[40] Oleguer Plana-Ripoll et al. “Analysis of mortality metrics associated with a comprehensive range of disorders in Denmark, 2000 to 2018: A population-based cohort study”. en. In: PLoS Med. 19.6 (June 2022), e1004023.

[41] F Privé et al. “Efficient analysis of large-scale genome-wide data with two R packages: bigstatsr and bigsnpr”. In: Bioinformatics 34.16 (Aug. 2018).

[42] Florian Privé, Hugues Aschard, and Michael GB Blum. “Efficient implementation of penalized regression for genetic risk prediction”. In: Genetics 212.1 (2019), pp. 65–74.

[43] Florian Privé et al. “Efficient toolkit implementing best practices for principal component analysis of population genetic data”. en. In: Bioinformatics 36.16 (May 2020), pp. 4449–4457.

[44] S L Pulit et al. “Meta-analysis of genome-wide association studies for body fat distribution in 694 649 individuals of European ancestry”. In: Hum. Mol. Genet. 28.1 (Jan. 2019).

[45] J M Robins. “Correction for non-compliance in equivalence trials”. en. In: Stat. Med. 17.3 (Feb. 1998), 269–302, discussion 387–9.

[46] P Rovira et al. “Shared genetic background between children and adults with attention deficit/hyperactivity disorder”. In: Neuropsychopharmacology 45.10 (Sept. 2020).

[47] Hon-Cheong So and Pak C Sham. “A unifying framework for evaluating the predictive power of genetic variants based on the level of heritability explained”. en. In: PLoS Genet. 6.12 (Dec. 2010), e1001230.

[48] James R Staley et al. “A comparison of Cox and logistic regression for use in genome-wide association studies of cohort and case-cohort design”. en. In: Eur. J. Hum. Genet. 25.7 (June 2017), pp. 854–862.

[49] Hamzah Syed, Andrea L Jorgensen, and Andrew P Morris. “Evaluation of methodology for the analysis of ‘time-to-event’ data in pharmacogenomic genome-wide association studies”. en. In: Pharmacogenomics 17.8 (June 2016), pp. 907–915.

[50] G Thorleifsson et al. “Genome-wide association yields new sequence variants at seven loci that associate with measures of obesity”. In: Nat. Genet. 41.1 (Jan. 2009).

[51] M Vujkovic et al. “Discovery of 318 new risk loci for type 2 diabetes and related vascular outcomes among 1.4 million participants in a multi-ancestry meta-analysis”. In: Nat. Genet. 52.7 (July 2020).

[52] Y Wu et al. “Multi-trait analysis for genome-wide association study of five psychiatric disorders”. In: Transl. Psychiatry 10.1 (June 2020).

[53] Jian Yang et al. “Advantages and pitfalls in the application of mixed-model association methods”. en. In: Nat. Genet. 46.2 (Feb. 2014), pp. 100–106.

[54] L Yengo et al. “Meta-analysis of genome-wide association studies for height and body mass index in 700000 individuals of European ancestry”. In: Hum. Mol. Genet. 27.20 (Oct. 2018).

[55] Noah Zaitlen et al. “Informed conditioning on clinical covariates increases power in case-control association studies”. en. In: PLoS Genet. 8.11 (Nov. 2012), e1003032.

[56] Noah Zaitlen et al. “Using extended genealogy to estimate components of heritability for 23 quantitative and dichotomous traits”. In: PLoS Genet. 9.5 (May 2013), e1003520.

[57] Z Zhu et al. “Shared genetic and experimental links between obesity-related traits and asthma subtypes in UK Biobank”. In: J. Allergy Clin. Immunol. 145.2 (Feb. 2020).

