## supplementalMaterial for "ADuLT: An efficient and robust time-to-event GWAS"

### 7 Supplemental Information

#### 7.1 Simulation Results

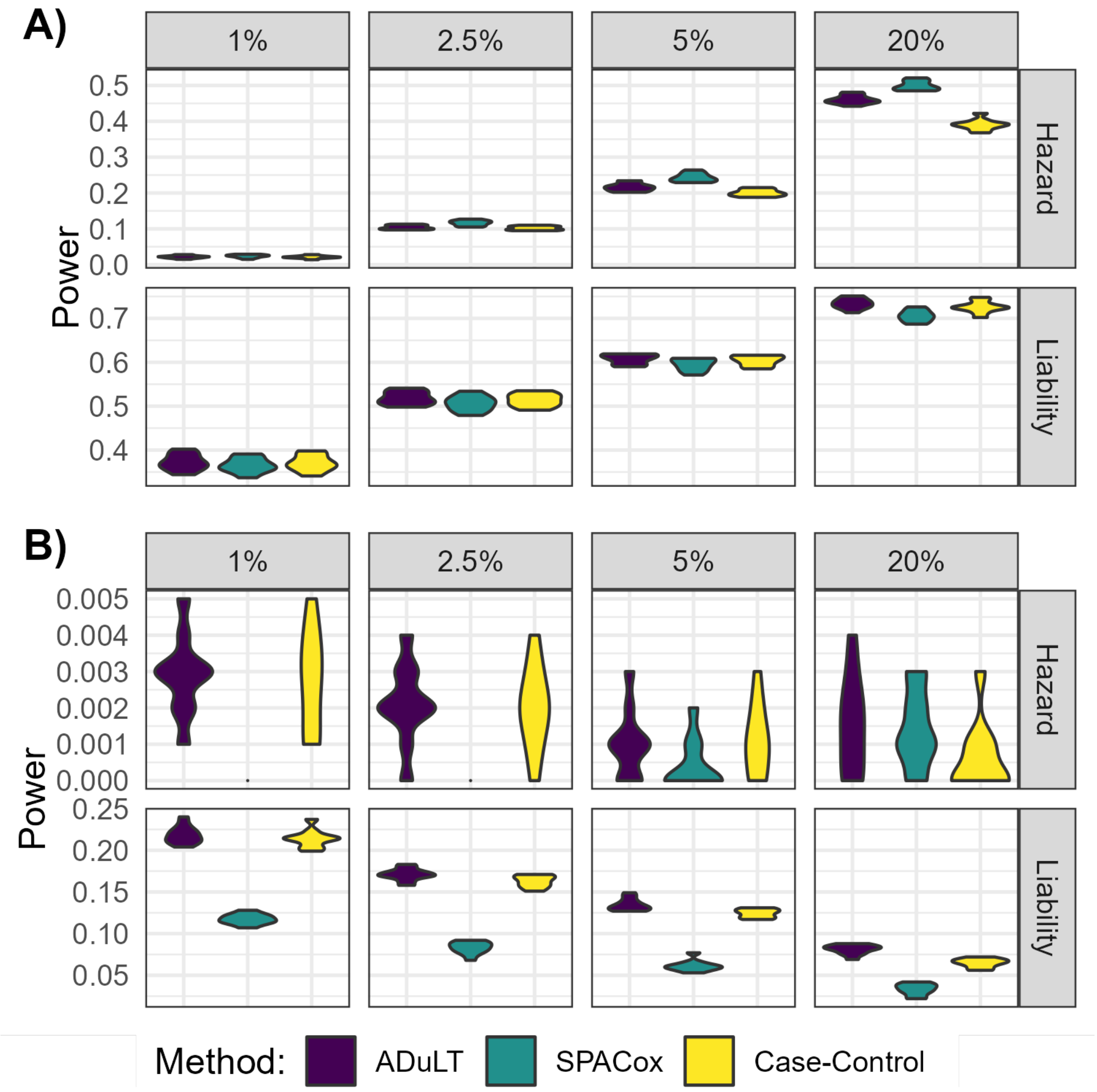


Figure S1: **Power simulation results with 1000 causal SNPs under both generative models and varying prevalences..** The power, i.e. the fraction of causal SNPs detected for each of the three methods, is shown for several prevalences, varying from 1% to 20%. **A)** The power of ADuLT, SPACox and case-control GWAS **without downsampling**. **B)** The power for the same three methods but **with downsampling**. When downsampling, the number of individuals is set to 20k, with 10k cases and 10k controls. This is done to assess performance in biobanks where cases have been ascertained.

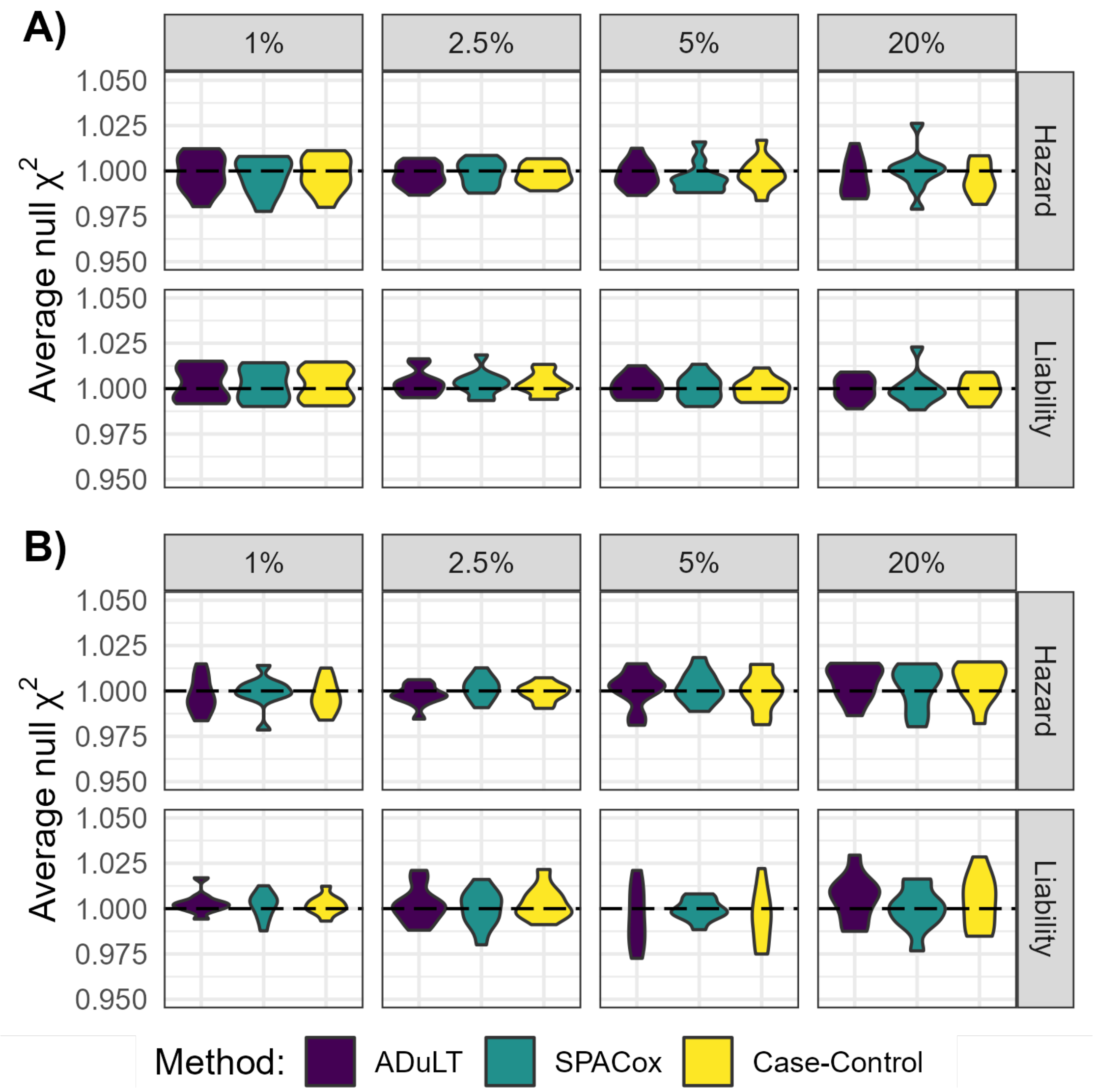


Figure S2: **Null SNPs -statistic simulation results with 250 causal SNPs under both generative models and varying prevalences.** The average null -statistic is shown for several prevalences, varying from 1% to 20%. **A)** The average null statistics for ADuLT, SPACox and case-control GWAS **without downsampling**. **B)** The average null statistics for the same three methods, but **with downsampling**. When downsampling, the number of individuals is set to 20k, with 10k cases and 10k controls. This is done to assess performance in biobanks where cases have been ascertained.

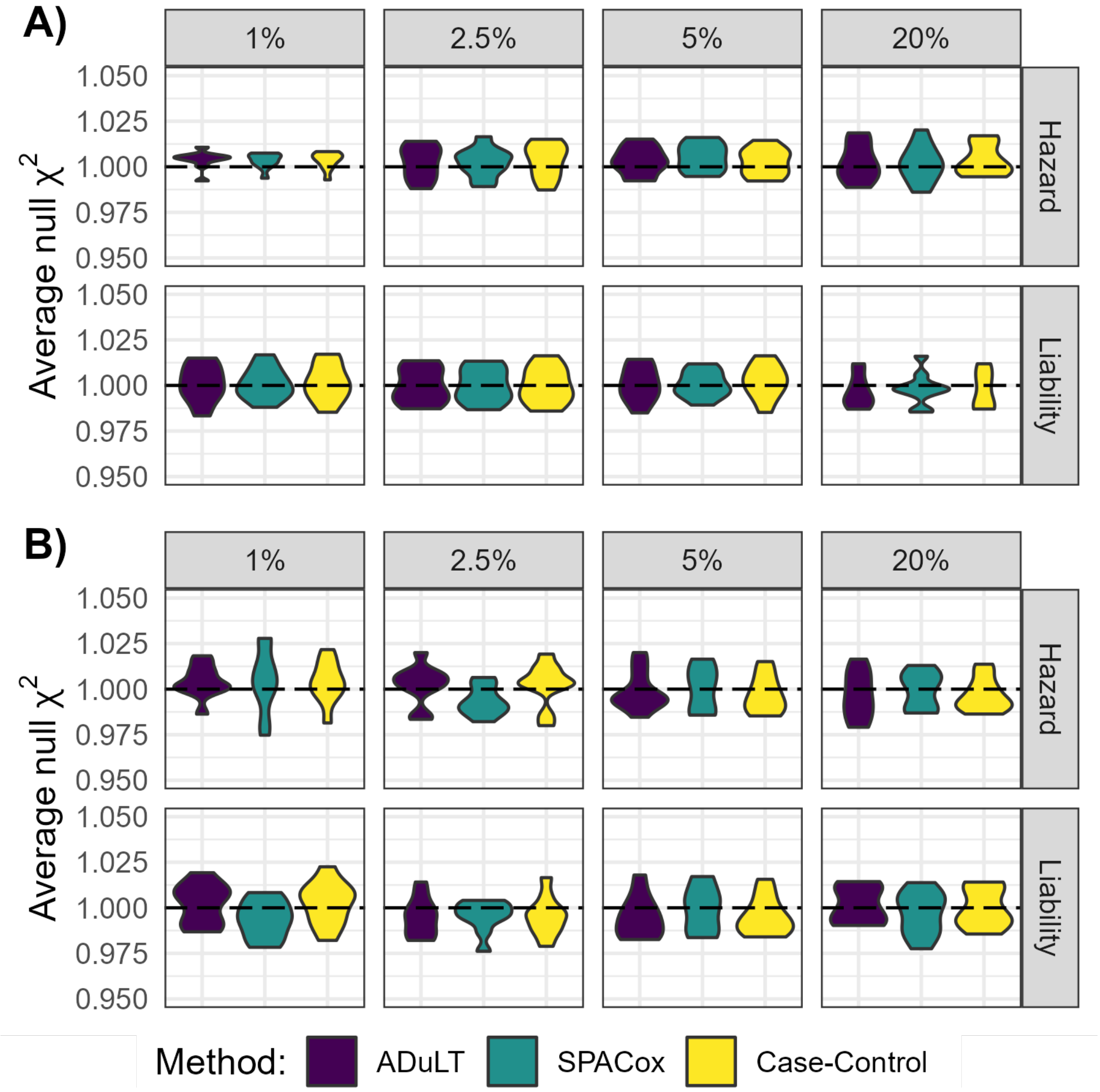


Figure S3: **Null SNPs -statistic simulation results with 1000 causal SNPs under both generative models and varying prevalences..** The average null -statistic is shown for several prevalences varying from 1% to 20%. **A)** The average null statistics for ADuLT, SPACox and case-control GWAS **without downsampling**. **B)** The average null statistics for the same three methods, but **with downsampling**. When downsampling, the number of individuals is set to 20k, with 10k cases and 10k controls. This is done to assess performance in biobanks where cases have been ascertained.

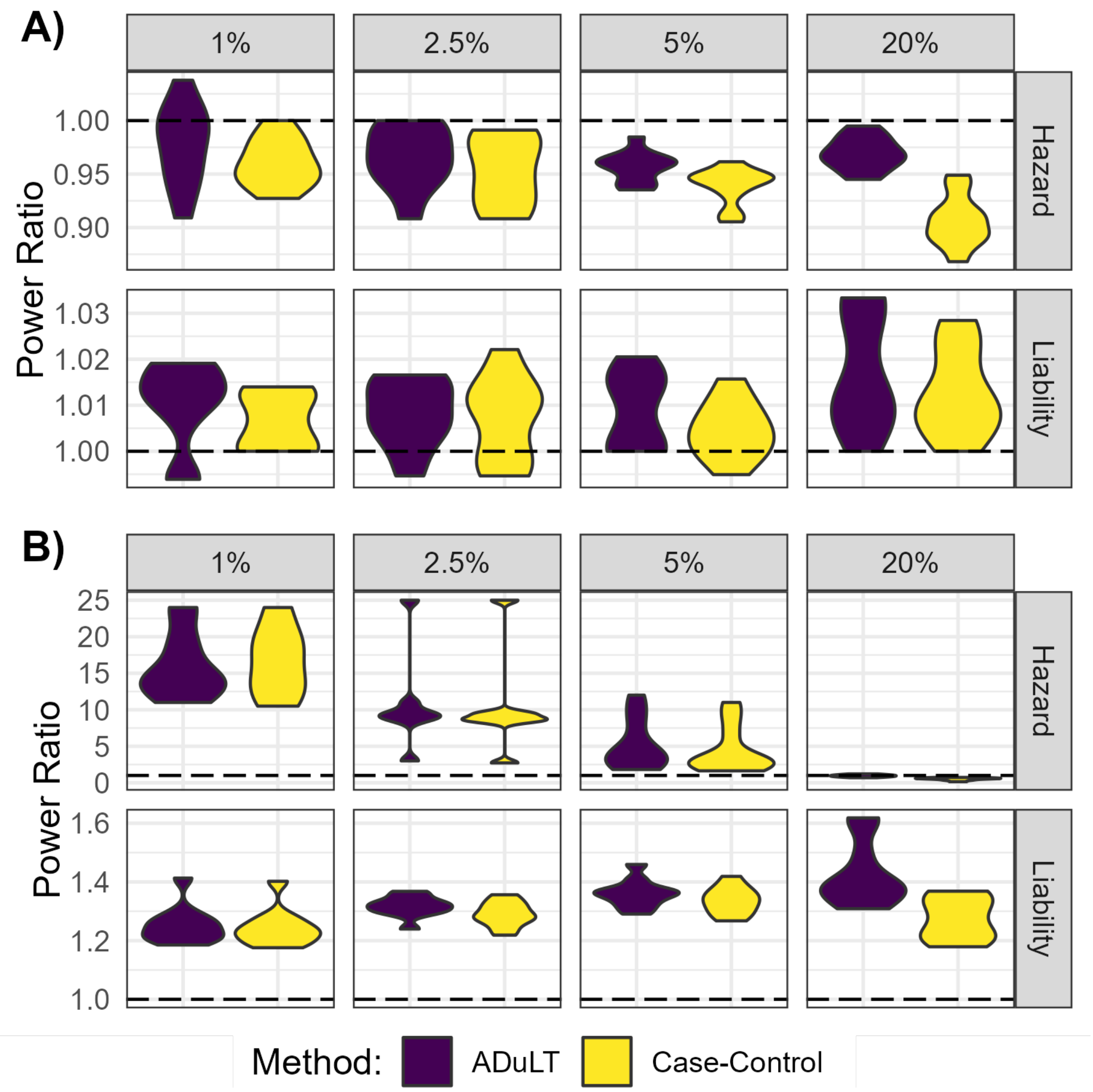


Figure S4: **Relative power ratio simulation results with 250 causal SNPs under two generative models and varying prevalences. .** The relative power is shown for several prevalences varying from 1% to 20%. SPACox is set to be the baseline method, and the ratio between the observed power of ADuLT GWAS as well as case-control GWAS is computed. The two plots show **A)** the power ratio **without downsampling**. **B)** the power ratio **with downsampling**. When downsampling, the number of individuals is set to 20k, with 10k cases and 10k controls. This is done to assess performance in biobanks where cases have been ascertained.

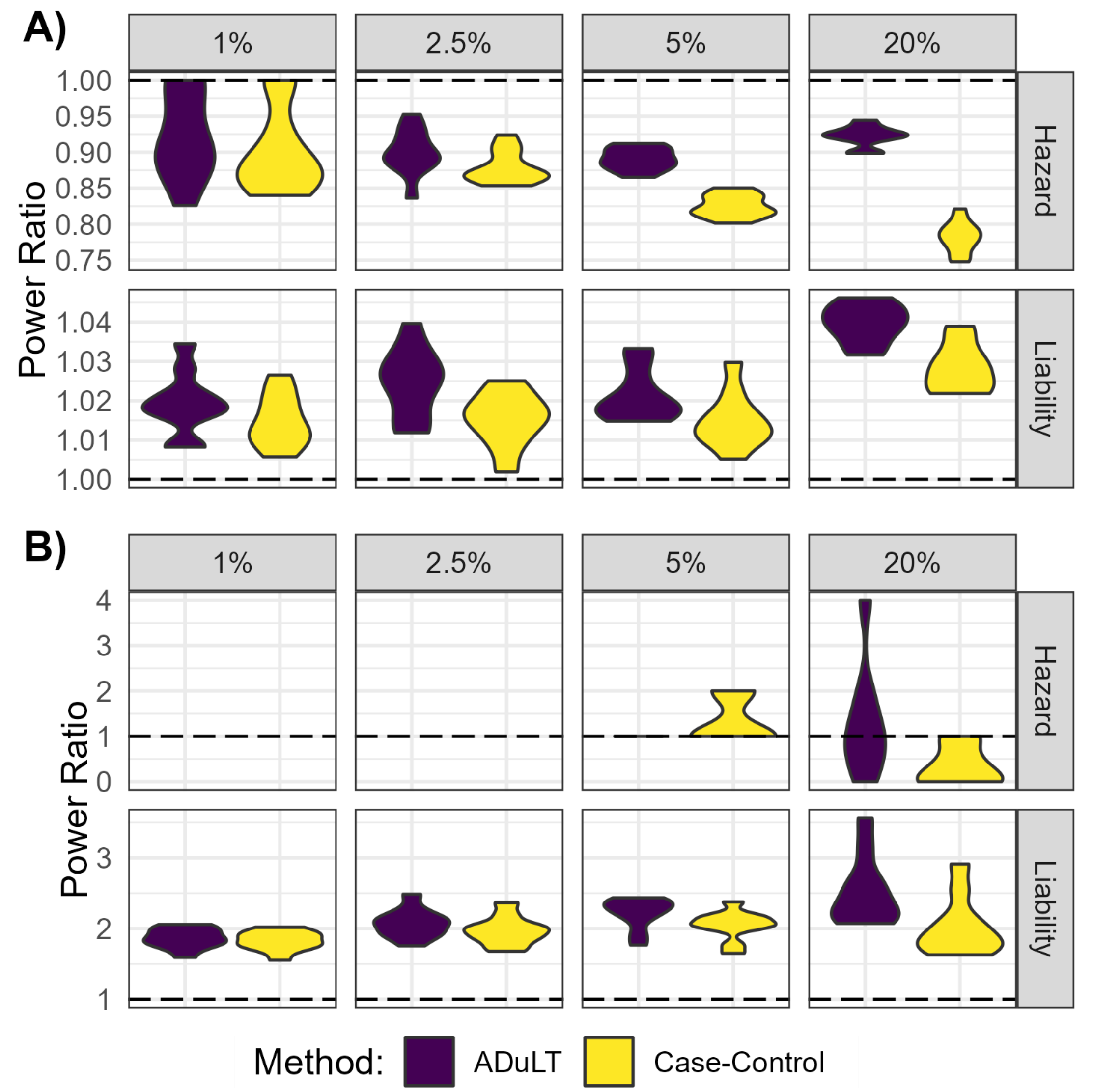


Figure S5: **Relative power ratio simulation results with 1000 causal SNPs and under two generative models and varying prevalences..** The relative power is shown for several prevalences varying from 1% to 20%. SPACox is set to be the baseline method, and the ratio between the observed power of the ADuLT GWAS as well as case-control GWAS is computed. The two plots show **A)** the power ratio **without downsampling**. **B)** the power ratio **with downsampling**. When downsampling, the number of individuals is set to 20k, with 10k cases and 10k controls. This is done to assess performance in biobanks where cases have been ascertained.

#### 7.2 iPSYCH Results

##### 7.2.1 ADHD

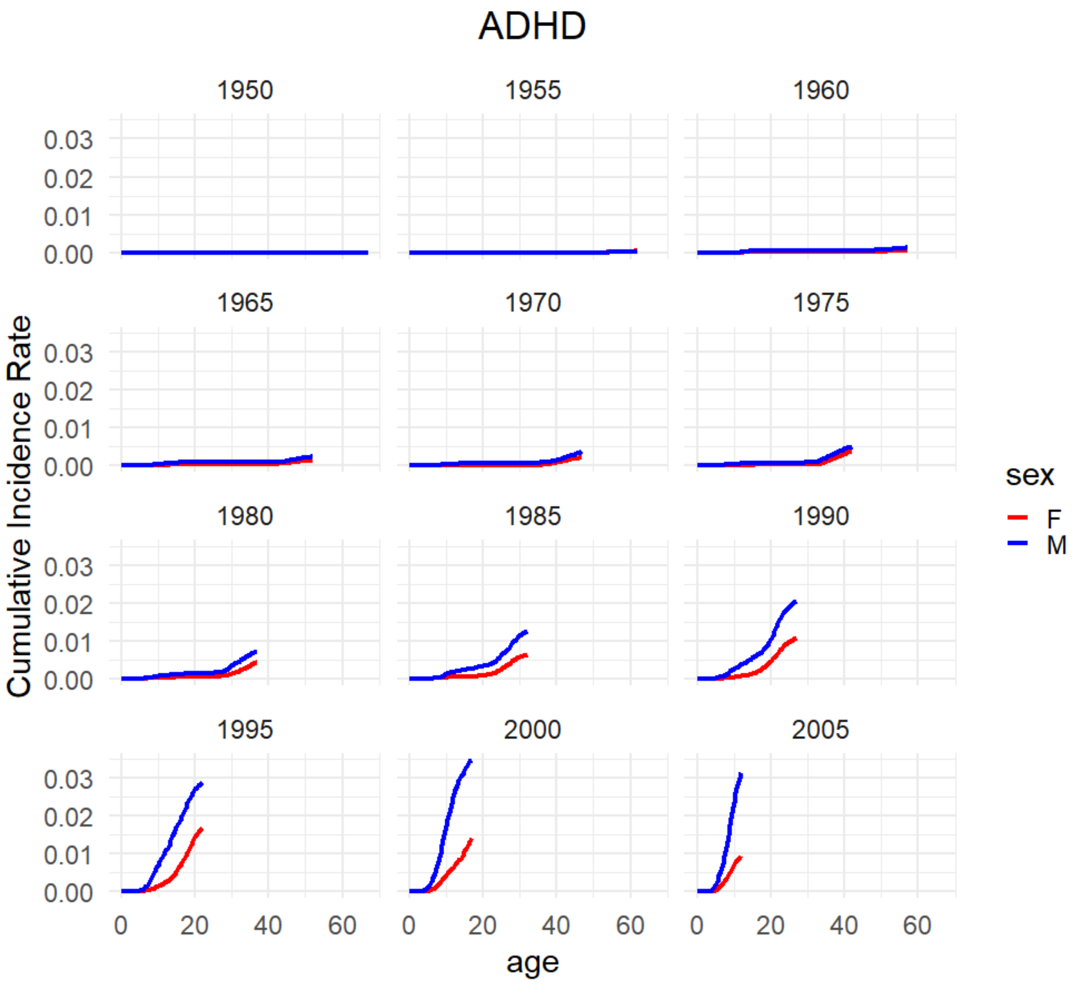


Figure S6: **Cumulative incidence rates for ADHD.** Cumulative incidence rates for ADHD in the Danish registers. The cumulative incidence proportions are stratified by birth year and sex.

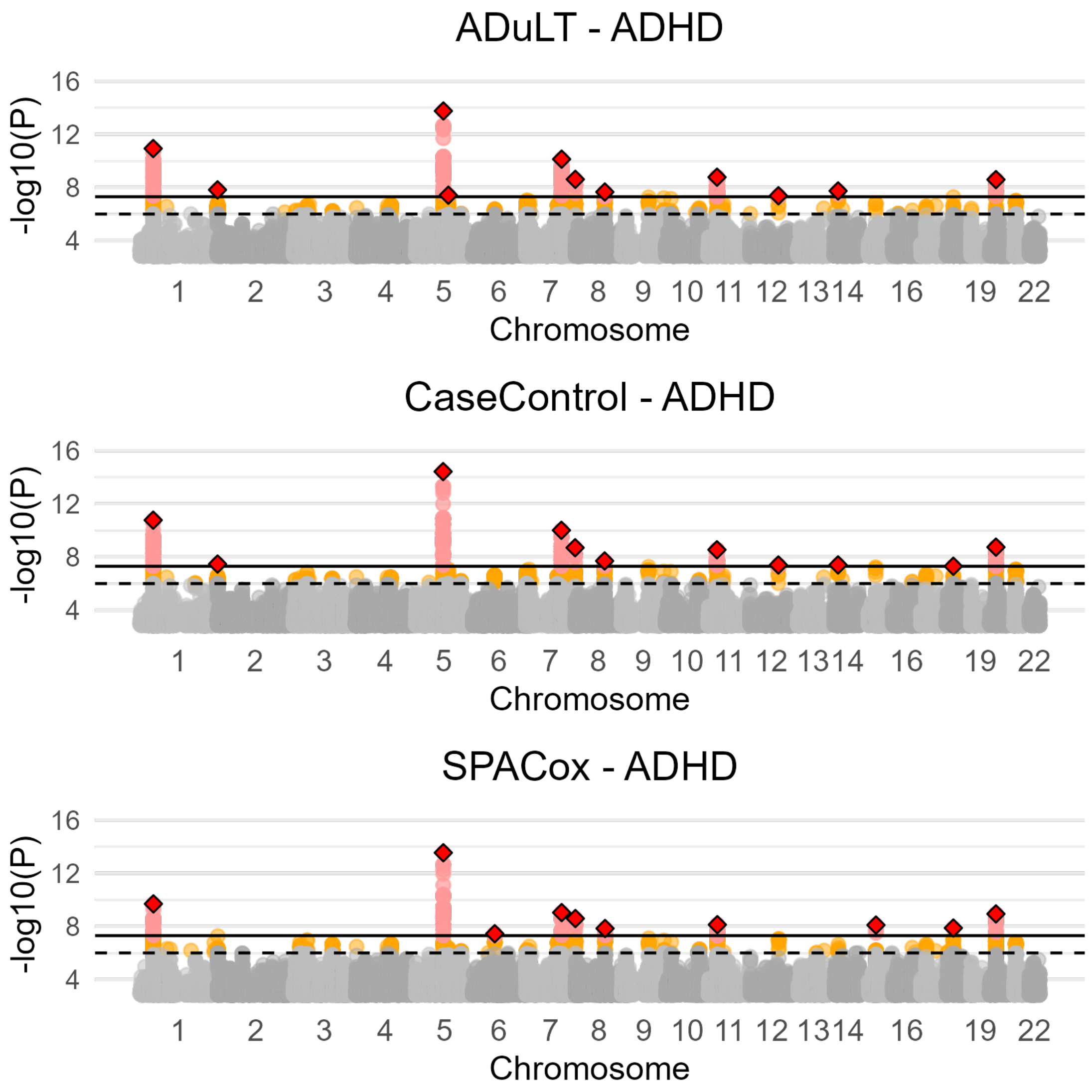


Figure S7: **Manhattan plots for SPACox as well as for a GWAS with ADuLT and case-control status as the outcome for ADHD with age as a covariate for all phenotypes..** Manhattan plots for ADHD based on the ADuLT GWAS, case-control GWAS and SPACox. The orange dots indicate suggestive SNPs with a p-value threshold of $5 \times 10^{-6}$. The red dots correspond to genome-wide significant SNPs with a p-value threshold of $5 \times 10^{-8}$. The diamonds correspond to the lowest p value LD clumped SNP in a 500k base pair window with a $r^2 = 0.1$ threshold.

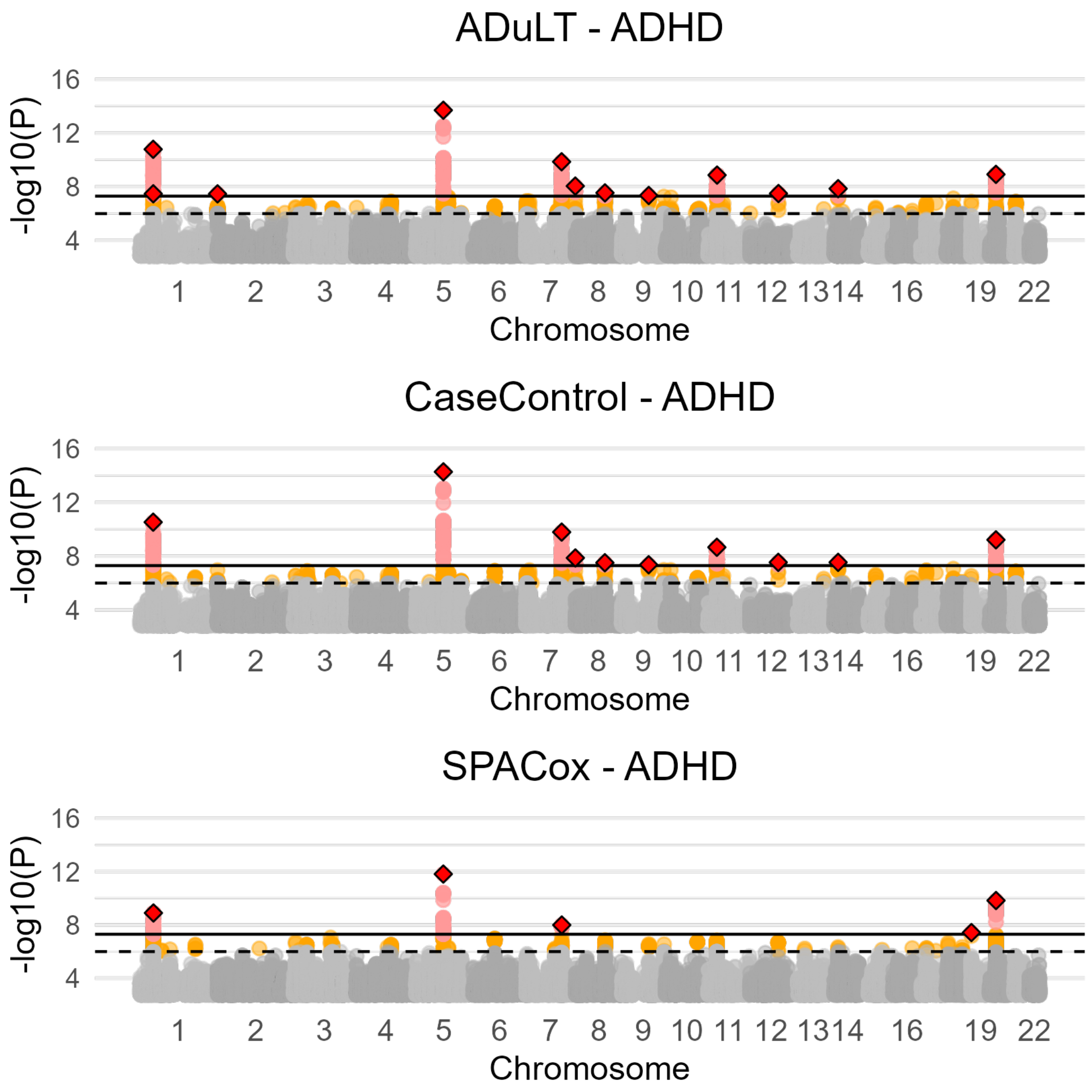


Figure S8: **Manhattan plots for ADuLT, case-control status, and SPACox of ADHD without age as a covariate for all phenotypes..** Manhattan plots for ADHD using the three methods. The orange dots indicate suggestive SNPs with a p-value threshold of $5 \times 10^{-6}$. The red dots correspond to genome-wide significant SNPs with a p-value threshold of $5 \times 10^{-8}$. The diamonds correspond to the lowest p value LD clumped SNP in a 500k base pair window with a $r^2 = 0.1$ threshold.

###### 7.2.2 Autism

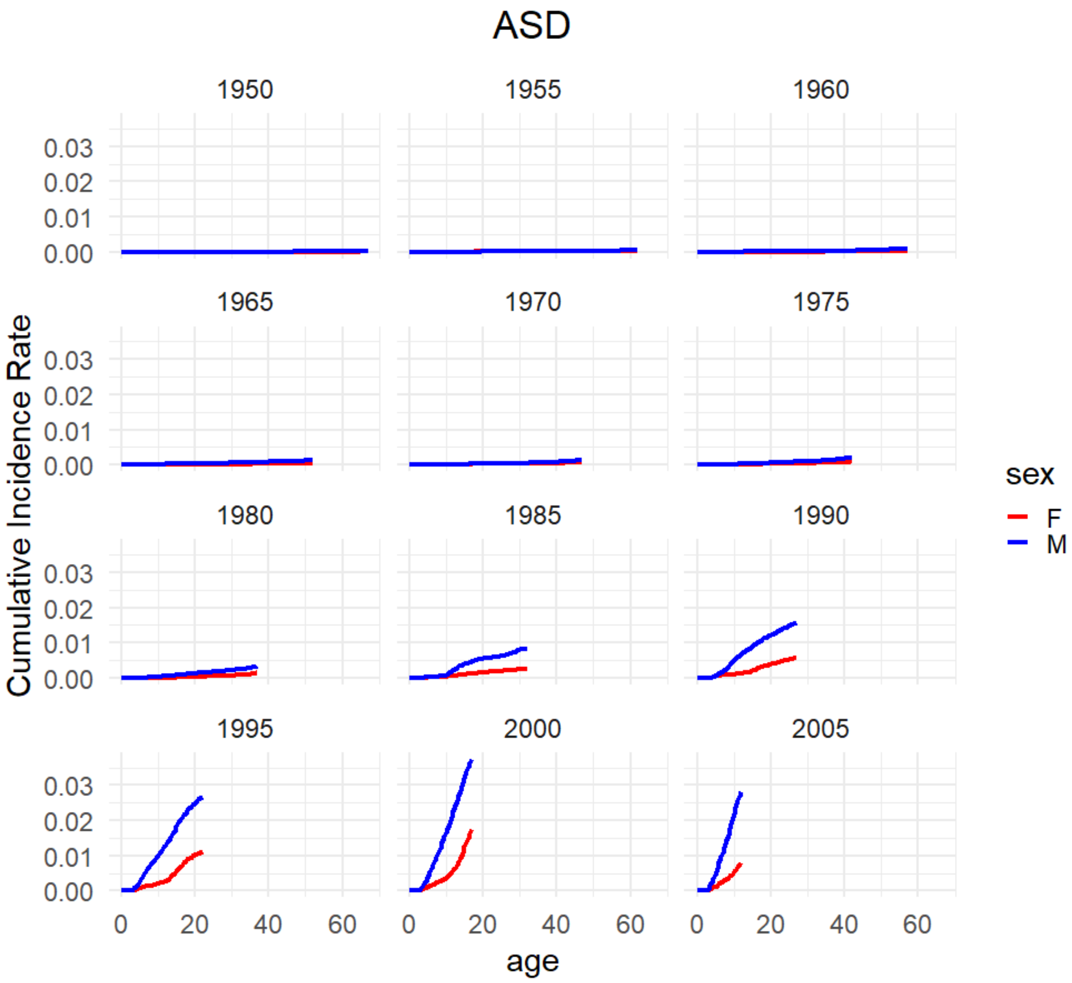


Figure S9: **Cumulative incidence rates for Autism.** Cumulative incidence rates for autism in the Danish registers. The cumulative incidence proportions are stratified by birth year and sex.

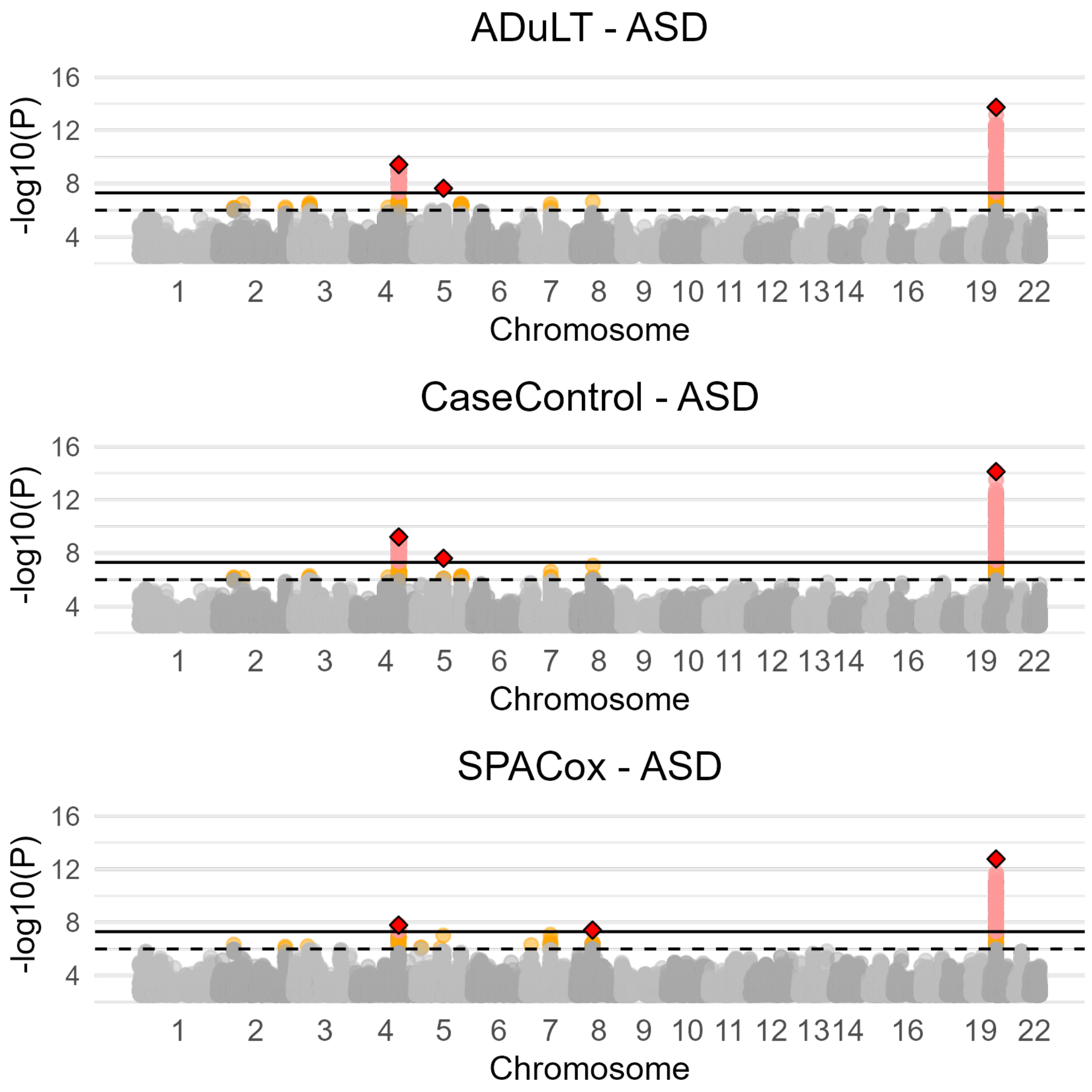


Figure S10: **Manhattan plots for ADuLT, case-control status, and SPACox of autism with age as a covariate for all phenotypes..** Manhattan plots for autism using the three methods. The orange dots indicate suggestive SNPs with a p-value threshold of $5 \times 10^{-6}$. The red dots correspond to genome-wide significant SNPs with a p-value threshold of $5 \times 10^{-8}$. The diamonds correspond to the lowest p value LD clumped SNP in a 500k base pair window with a $r^2 = 0.1$ threshold.

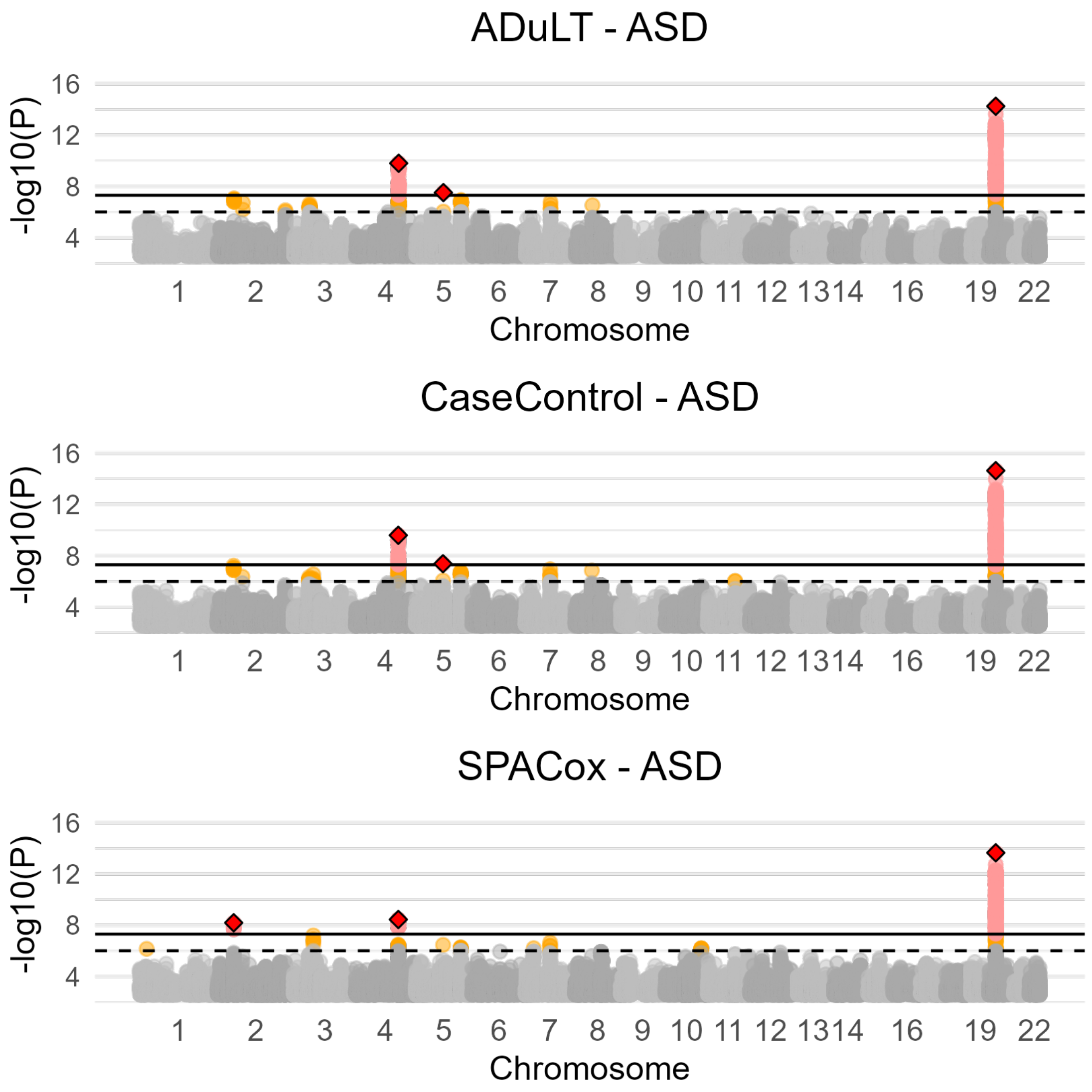


Figure S11: **Manhattan plots for ADuLT, case-control status, and SPACox of autism without age as a covariate for all phenotypes..** Manhattan plots for autism using the three methods. The orange dots indicate suggestive SNPs with a p-value threshold of $5 \times 10^{-6}$. The red dots correspond to genome-wide significant SNPs with a p-value threshold of $5 \times 10^{-8}$. The diamonds correspond to the lowest p value LD clumped SNP in a 500k base pair window with a $r^2 = 0.1$ threshold.

##### 7.2.3 Depression

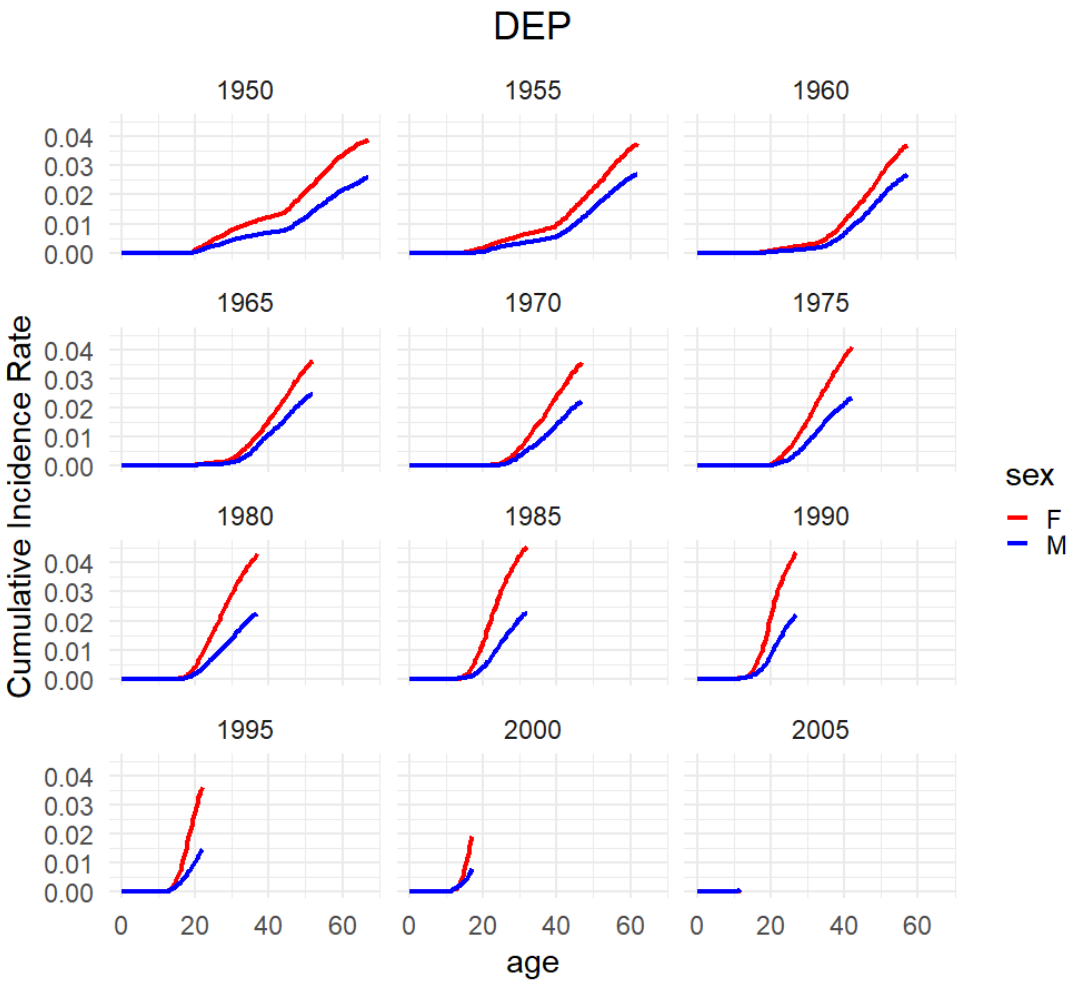


Figure S12: **Cumulative incidence rates for Depression.** Cumulative incidence rates for depression in the Danish registers. The cumulative incidence proportions are stratifiedby birth year and sex.

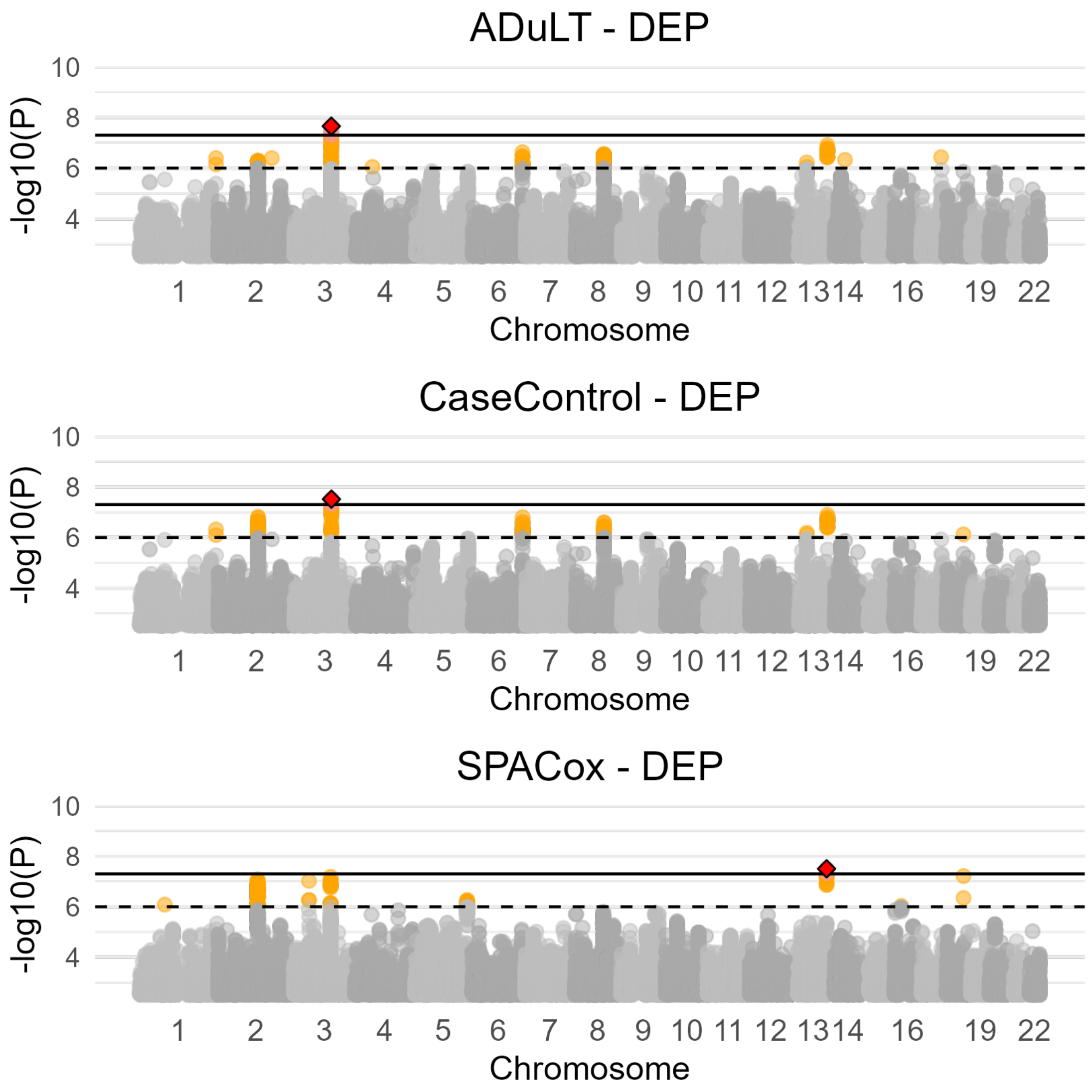


Figure S13: **Manhattan plots for ADuLT, case-control status, and SPACox of depression with age as a covariate for all phenotypes..** Manhattan plots for depression using the three methods. The orange dots indicate suggestive SNPs with a p-value threshold of $5 \times 10^{-6}$. The red dots correspond to genome-wide significant SNPs with a p-value threshold of $5 \times 10^{-8}$. The diamonds correspond to the lowest p value LD clumped SNP in a 500k base pair window with a $r^2 = 0.1$ threshold.

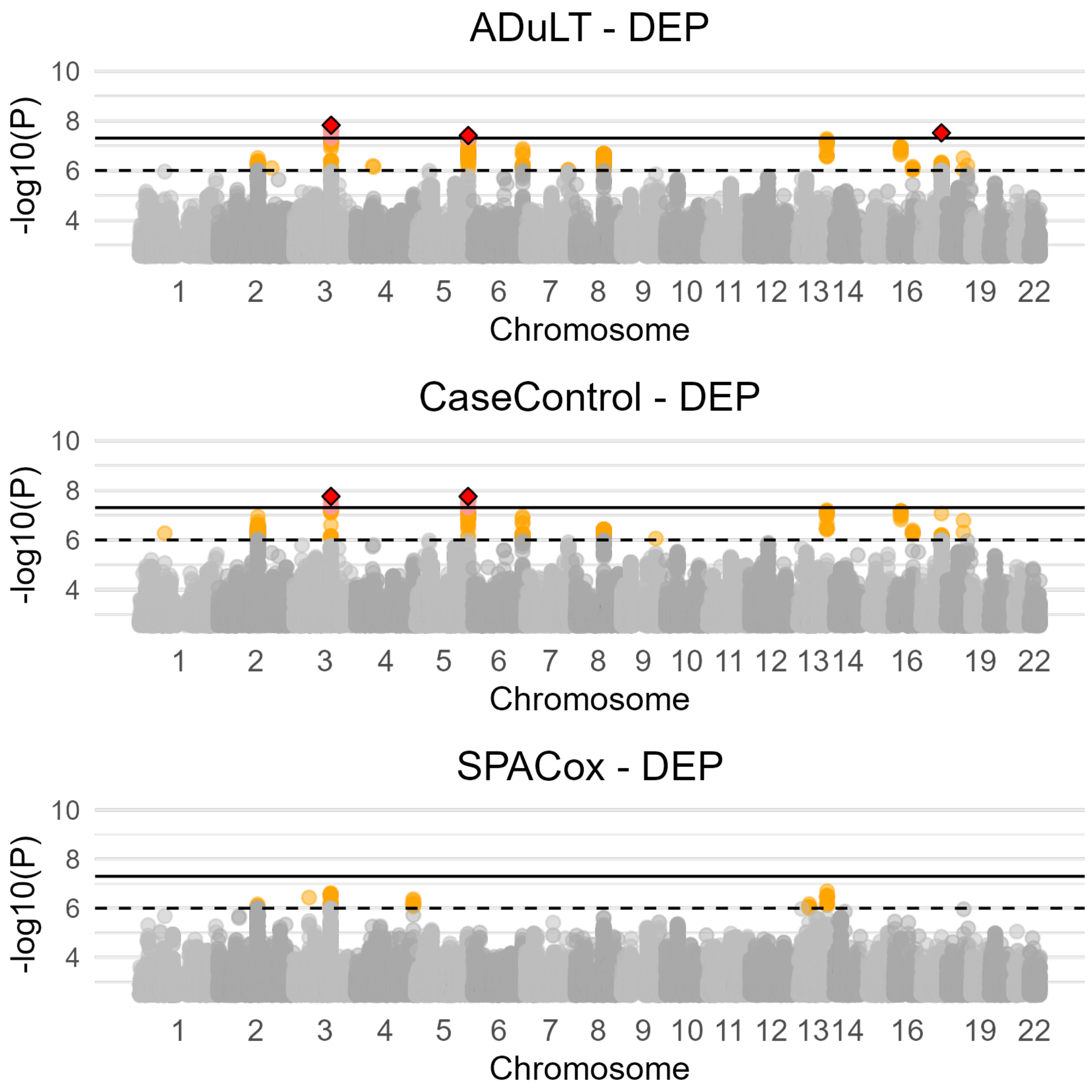


Figure S14: **Manhattan plots for ADuLT, case-control status, and SPACox of depression without age as a covariate for all phenotypes..** Manhattan plots for depression using all three methods. The orange dots indicate suggestive SNPs with a p-value threshold of $5 \times 10^{-6}$. The red dots correspond to genome-wide significant SNPs with a p-value threshold of $5 \times 10^{-8}$. The diamonds correspond to the lowest p value LD clumped SNP in a 500k base pair window with a $r^2 = 0.1$ threshold.

##### 7.2.4 Schizophrenia

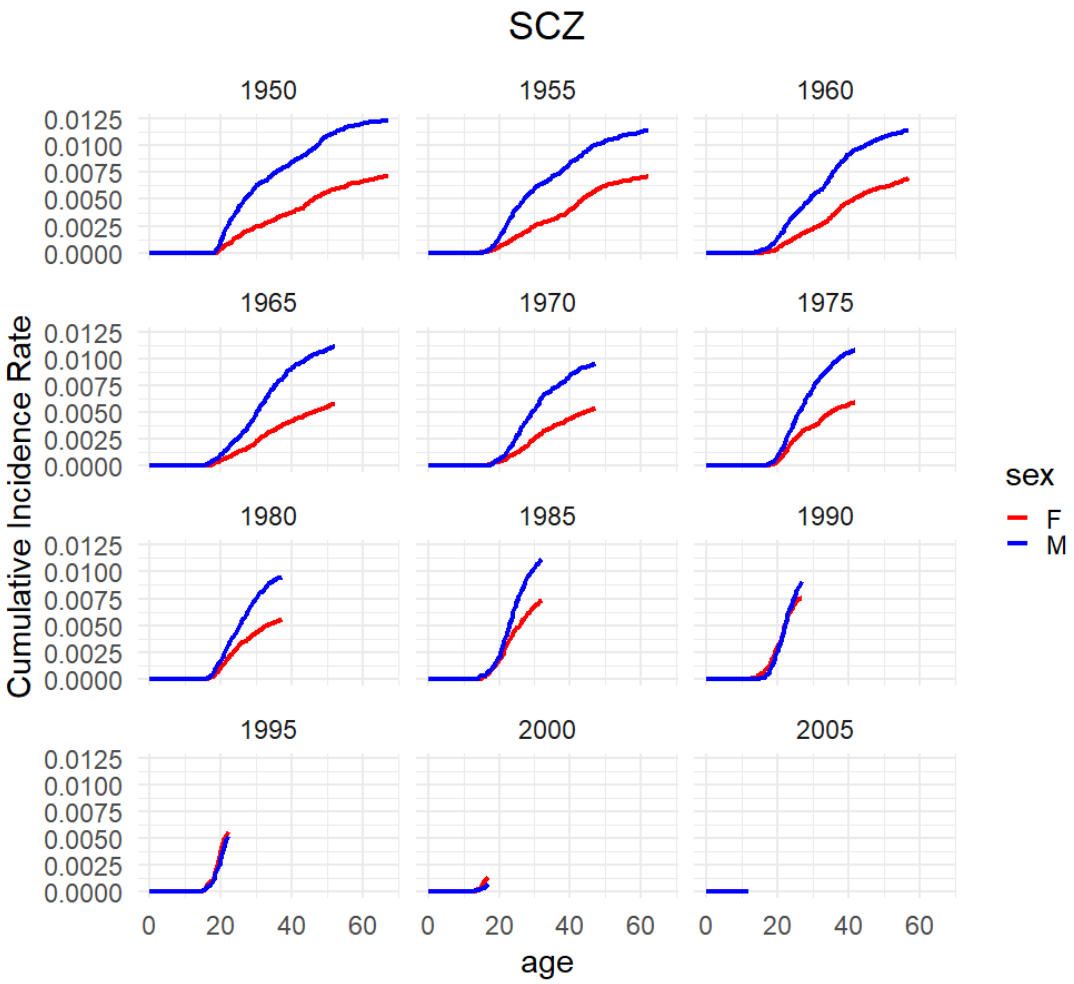


Figure S15: **Cumulative incidence rates for Schizophrenia.** Cumulative incidence rates for schizophrenia in the Danish registers. The cumulative incidence proportions are stratified by birth year and sex.

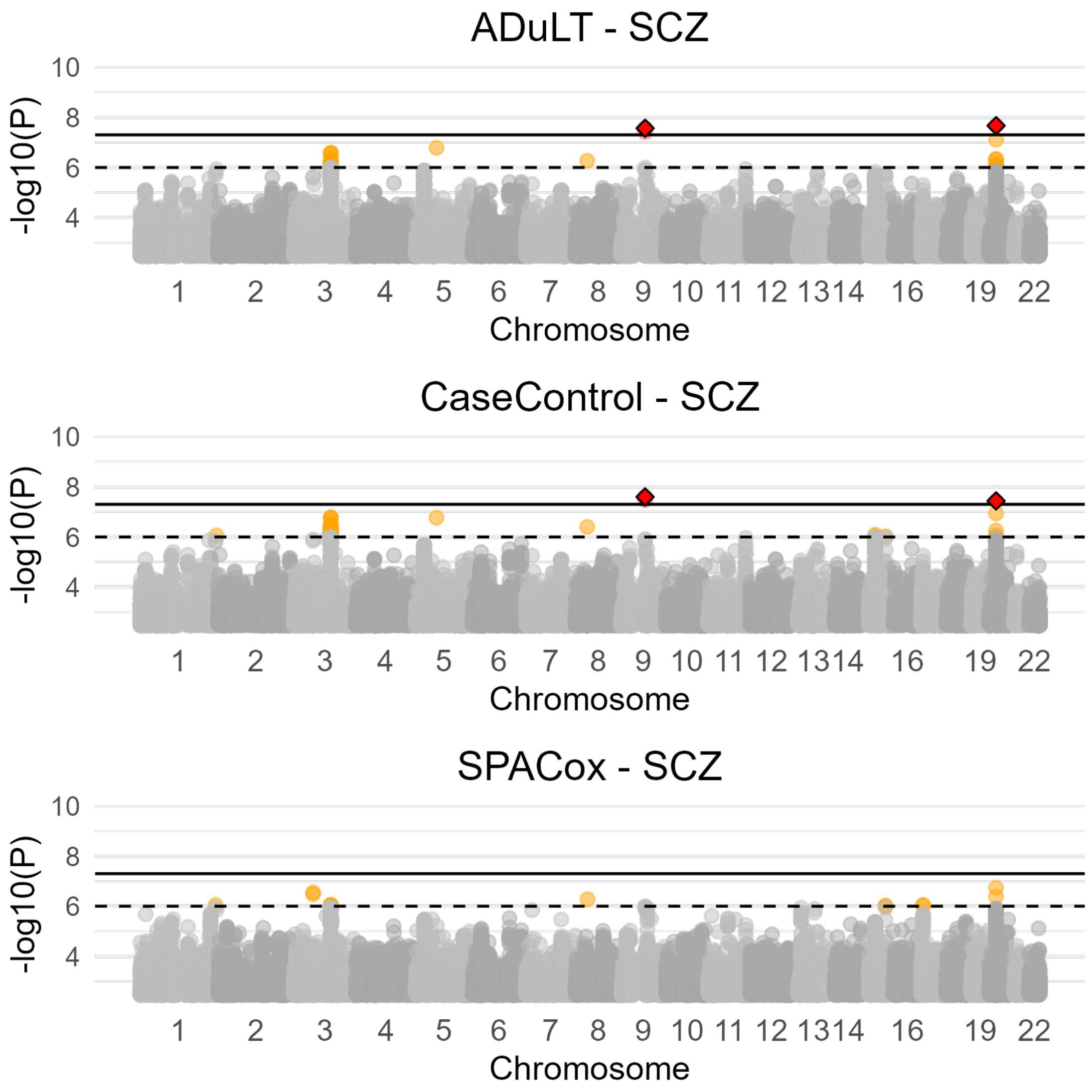


Figure S16: **Manhattan plots for ADuLT, case-control status, and SPACox of schizophrenia with age as a covariate for all phenotypes..** Manhattan plots for schizophrenia using the three methods. The orange dots indicate suggestive SNPs with a p-value threshold of $5 \times 10^{-6}$. The red dots correspond to genome-wide significant SNPs with a p-value threshold of $5 \times 10^{-8}$. The diamonds correspond to the lowest p value LD clumped SNP in a 500k base pair window with a $r^2 = 0.1$ threshold.

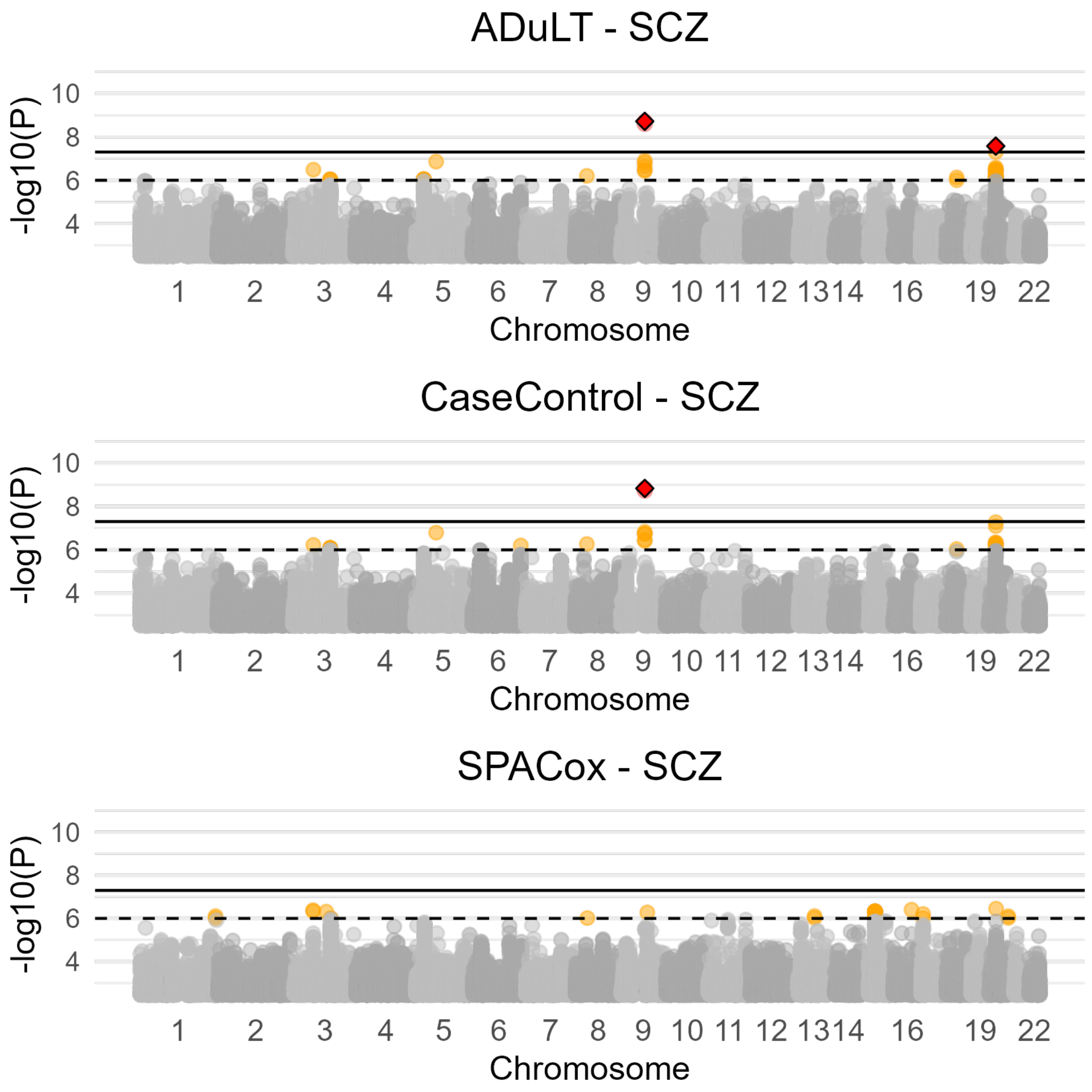


Figure S17: **Manhattan plots for ADuLT, case-control status, and SPACox of schizophrenia without age as a covariate for all phenotypes..** Manhattan plots for schizophrenia using the three methods. The orange dots indicate suggestive SNPs with a p-value threshold of $5 \times 10^{-6}$. The red dots correspond to genome-wide significant SNPs with a p-value threshold of $5 \times 10^{-8}$. The diamonds correspond to the lowest p value LD clumped SNP in a 500k base pair window with a $r^2 = 0.1$ threshold.

#### 7.3 Tables

Table S1: **Excel file with summary information from the simulations, containing information such as power, null $\chi^2$-statistics, false positive rates, etc.** All summary information from the simulations has been combined into this Excel file, and it contains all relevant information in a summary format, e.g. power, null $\chi^2$-statistics, false positive rates, etc. All information is available for each parameter setup and the parameters consist of the generative model, prevalence, phenotype, number of causal SNPs, and whether downsampling was applied or not. Each simulation setup has 10 replications, and information is available for each iteration.

| | ADHD | Autism | Depression | Schizophrenia |
|---|---|---|---|---|
| Control | 36548 | 36741 | 36368 | 36921 |
| Case | 21738 | 18235 | 27507 | 11602 |

Table S2: **Table including the number of individuals used for each iPSYCH disorders.** The number of cases and controls used in the GWAS for each iPSYCH phenotype. Numbers are shown after filtering for relatedness and restricting to a group of individuals with European ancestry.

| Variant ID | Chromosome: Position (hg38) | Effect size (SE) | ADuLT p-value (-log10(P)) | Nearest gene | Selected previously reported associations |
|---|---|---|---|---|---|
| rs11210887 | 1:44076019 | 0.0331(0.0049) | 10.8 | PTPRF | smoking initiation, educational attainment[27, 29] |
| rs11210887 | 1:44076019 | 0.0331(0.0049) | 10.8 | PTPRF | smoking initiation, educational attainment[27, 29] |
| rs4660756 | 1:44383914 | 0.0284(0.0051) | 7.48 | ST3GAL3 | educational attainment, ADHD [36, 52] |
| rs7563362 | 2:620297 | -0.0361(0.0065) | 7.47 | LINC01875, TMEM18 | Type 2 diabetes, BMI [50, 51] |
| rs4916723 | 5:87854395 | -0.0359(0.0047) | 13.7 | LINC00461 | ADHD, Educational Attainment, BMI [52, 27, 57] |
| rs12705966 | 7:114248851 | 0.0334(0.0052) | 9.86 | - | - |
| rs13236619 | 7:157827565 | -0.0263(0.0046) | 8.05 | - | - |
| rs72673548 | 8:93292844 | -0.0423(0.0076) | 7.55 | - | - |
| rs12346733 | 9:86727865 | -0.0256(0.0047) | 7.35 | - | - |
| rs57806515 | 11:28628549 | 0.0282(0.0047) | 8.87 | - | - |
| rs704061 | 12:89771903 | -0.0252(0.0046) | 7.49 | DUSP6, POC1B | ADHD, Educational Attainment, BMI [52, 27, 44] |
| rs4261436 | 14:33299482 | -0.0257(0.0045) | 7.86 | AKAP6 | Educational attainment, BMI, Type 2 diabetes[27, 57, 31] |
| rs4813421 | 20:21258053 | 0.0305(0.005) | 8.92 | - | - |

Table S3: **LD clumped genome-wide significant SNPs based on the ADuLT phenotype for ADHD in iPSYCH.** SNPs are ordered by chromosome and location. The table contains the rsID, chromosome, location (bp), effect size, standard error, and p-value for each SNP. Information on the closest gene and some selected previous associations for each SNP are included as well.

| Variant ID | Chromosome: Position (hg38) | Effect size (SE) | ADuLT p-value (-log10(P)) | Nearest gene | Selected previously reported associations |
|---|---|---|---|---|---|
| rs8085882 | 18:22743899 | 0.0156(0.0029) | 7.30 | ZNF521 | education attainment, smoking initiation[27, 29] |

Table S4: **LD clumped genome-wide significant SNPs for ADHD that are unique to Case-Control status in iPSYCH.** The table contains the rsID, chromosome, location (bp), effect size, standard error, and p-value for each SNP. Information on the closest gene and some selected previous associations for each SNP are also included.
